# Risk of Adverse Maternal and Fetal Outcomes Associated with COVID-19 Variants of Concern: A Sequential Prospective Meta-Analysis

**DOI:** 10.1101/2023.04.03.23287260

**Authors:** Fouzia Farooq, Erin Oakley, Daniel Kerchner, Jenny Yeon Hee Kim, Victor Akelo, Beth A. Tippett Barr, Elisa Bevilacqua, Nabal Bracero, Maria del Mar Gil, Camille Delgado-López, Guillaume Favre, Irene Fernandez Buhigas, Hiu Yu Hillary Leung, Valentina Laurita Longo, Alice Panchaud, Liona C. Poon, Raigam Jafet Martinez-Portilla, Miguel Valencia-Prado, James M. Tielsch, Emily R. Smith, the COVI-PREG International Registry, the PMA collaborators

**Affiliations:** Department of Global Health, Milken Institute School of Public Health, The George Washington University, Washington, DC, USA; George Washington University Libraries, The George Washington University, Washington, DC, USA; Department of Biostatistics & Bioinformatics, Milken Institute School of Public Health, The George Washington University, Washington, DC, USA; US Centers for Disease Control and Prevention, Kisumu, Kenya; Nyanja Health Research Institute, Salima, Malawi; Department of Women and Child Health, Women Health Area, Fondazione Policlinico Universitario Agostino Gemelli, IRCCS, Rome, Italy; University of Puerto Rico School of Medicine, Department of Obstetrics and Gynecology, Obstetrics and Gynecology (PROGyn), Puerto Rico; Department of Obstetrics and Gynecology, Hospital Universitario de Torrejón, Torrejón de Ardoz, Madrid, Spain; School of Medicine, Universidad Francisco de Vitoria, Pozuelo de Alarcón, Madrid, Spain; Surveillance for Emerging Threats to Mothers and Babies, Puerto Rico Department of Health, San Juan, Puerto Rico; Materno-fetal and Obstetrics Research Unit, Department “Femme-Mère-Enfant”, University Hospital, Lausanne, Switzerland; Catholic University of Sacred Heart, Rome, Italy; Institute of Primary Health Care (BIHAM), University of Bern, Bern, Switzerland; Department of Obstetrics and Gynecology, The Chinese University of Hong Kong, Hong Kong; Clinical Research Branch, National Institute of Perinatology, Mexico City, Mexico; Children with Special Medical Needs Division, Puerto Rico Department of Health, San Juan, Puerto Rico; Department of Exercise and Nutrition Sciences, Milken Institute School of Public Health, The George Washington University, Washington, DC, USA; Kenya Medical Research Institute, Center for Global Health Research [KEMRI-CGHR], Kisumu, Kenya; KEMRI-CGHR, Kisumu, Kenya; Department of Obstetrics and Gynecology, Pontoise Hospital, Pontoise, France; Obstetrics Division, Department of Pediatrics, Gynecology, and Obstetrics Geneva University Hospitals & Faculty of Medicine, University of Geneva, Geneva, Switzerland; Department of Obstetrics and Gynecology, Poissy-Saint Germain Hospital, Poissy, France; Department of Obstetrics, Basel University Hospital, Basel, Switzerland; Department of Obstetrics and Gynaecology, Queen Elizabeth Hospital, Hong Kong SAR, China; Department of Obstetrics and Gynaecology, United Christian Hospital, Hong Kong SAR, China; Department of Obstetrics and Gynaecology, Princess Margaret Hospital, Hong Kong SAR, China; Department of Obstetrics and Gynaecology, Tuen Mun Hospital, Hong Kong SAR, China; Department of Obstetrics and Gynaecology, Kwong Wah Hospital, Hong Kong SAR, China; Department of Women’s and Child Health Sciences and Public Health, IRCCS A. Gemelli University Polyclinic Foundation, Rome, Italy

## Abstract

**Introduction:** The main objective of this study is to conduct an individual patient data meta-analysis with collaborators from various countries to identify SARS-CoV-2 variants of concern associated with adverse maternal and neonatal outcomes.

**Methods:** Eligible studies included registries and single- or multi-site cohort studies that recruited pregnant and recently postpartum women with confirmed COVID-19. Studies must have enrolled at least 25 women within a defined catchment area. Studies also had to have data that overlapped more than a single COVID-19 variant time period. We invited principal investigators already participating in an ongoing sequential, prospective meta-analysis of perinatal COVID-19. Investigators shared individual patient data (IPD) with the technical team for review and analysis. We examined 31 outcomes related to: i) COVID-19 severity (n=5); ii) maternal morbidities including adverse birth outcomes (n=14); iii) fetal and neonatal morbidity and mortality (n=5) and iv) adverse birth outcomes (n=8). SARS-CoV-2 strains that have been identified as variants of concern (VOC) by the WHO were analyzed using the publicly available strain frequency data by Nextstrain.org and strains were classified as dominant when they were more than half of sequences in a given geographic area. We applied a 2-stage IPD meta-analytic framework to generate pooled relative risks, with 95% CI for each dominant variant and outcome pair when there were one or more studies with available data.

**Results:** Our data show that the Delta wave, compared to Omicron, was associated with a higher risk of all adverse COVID-19 severity outcomes in pregnancy including risk of hospitalization [RR 4.02 (95% CI 1.10, 14.69), n=1 study], risk of ICU admissions [RR 2.59 (95% CI 1.26, 5.30, n=3 studies], risk of critical care admission [RR 2.52 (95% CI 1.25, 5.08, n=3 studies], risk of needing ventilation [RR 3.96 (95% CI 1.47, 10.71), n=3 studies] and risk of pneumonia [RR 6.73 (95% CI 2.17, 20.90), n=3 studies]. The majority of maternal morbidity and mortality indicators were not at increased risk during any of the COVID-19 variant waves except hemorrhage, any Cesarean section, intrapartum Cesarean section and maternal composite outcome, although data was limited. Risk of fetal and neonatal morbidity and mortality did not show significant increases in risks during any of the COVID-19 waves except stillbirth and perinatal death during the Delta wave ([RR 4.84 (95% CI 1.37, 17.05, n=3 studies], [RR 6.03 (95%CI 1.63, 22.34), n=3 studies], respectively) when compared to the Pre-alpha wave. Adverse birth outcomes including very low birthweight and very preterm birth also showed increased risks during the Delta wave compared to the Pre-alpha wave.

**Discussion:** During periods of Delta strain predominance, all COVID-19 severity outcomes were more severe among pregnant women, compared to periods when other COVID-19 strains predominated. In addition, there are limited data comparing the impact of different variants on pregnancy outcomes. This highlights the importance of ongoing genomic surveillance among special populations.

## Introduction

The current Coronavirus disease 2019 (COVID-19) pandemic continues to affect all individuals across the globe. Although this single-stranded RNA virus of the *Coronaviridae* family named the severe acute respiratory syndrome coronavirus-2 (SARS-CoV-2)^1^ has been investigated in depth over the past two years, facets of its impact remain yet to be unraveled, particularly in sub-populations and under-resourced locations.

One population warranting further investigation is that of pregnant women and pregnant people^1a^, who are known to be at risk for more severe disease and more likely to require respiratory support upon infection.^2–6^ Our recent meta-analysis across 12 studies among 1942 pregnant women with COVID-19 compared to 11,194 non-COVID pregnancies showed that women with COVID-19 were 3.81 times more likely to require ICU admission, 15.23 times more likely to receive ventilation, and 7.68 times more likely to experience death compared to those without COVID-19.^7^ We also showed that neonates born to women with COVID-19 were 1.86 times more likely to be admitted to the neonatal care unit, 1.71 times more likely to be born preterm and 1.19 times more likely to be low birthweight compared to those without COVID-19. Other studies have also shown worse neonatal outcomes such as increased rates of preterm birth, admission to the neonatal intensive care unit (NICU), low birth weight, and fetal distress have also been associated with maternal SARS-CoV-2 infection.^3,8–10^

With a high mutation rate of the virus, the COVID-19 pandemic has been characterized by variant-specific epidemic waves and the severity of clinical outcomes has also differed by variant.^11–14^ Since the onset of pandemic, several variants of concern (VOCs) have been identified, including the Alpha (B.1.1.7), Beta (B.1.351), Gamma (P.1), Delta (B.1.617.2), Epsilon (B.1.427 and B.1.429), Eta (B.1.525), and Omicron (B.1.529, BA.1, BA.1.1, BA.2, BA.3, BA.4 and BA.5).^15^ A meta-analysis by Lin et al evaluating COVID-19 severity by variants showed the Beta and Delta variants as posing greater risk of hospitalization and mortality compared to the Alpha, Gamma, and wild-type (pre-Alpha) variants among the general population.^12^ A recent study conducted in the US by DeSisto et al reported a higher risk of stillbirths in mothers with COVID-19 during the period of Delta variant predominance^11^ Similarly, a UK study across 194 hospitals by Vousden et al suggested more severe maternal infection and worse pregnancy and perinatal outcomes in infected mothers during the Alpha and Delta dominance periods compared to the pre-Alpha dominance period.^4^ Another recent study by Favre et al compared maternal and perinatal outcomes across three variant periods (pre-Delta, Delta, and Omicron) in France and Switzerland and showed that the Delta period is associated with higher risk of adverse maternal outcomes compared to the pre-Delta period and the Omicron period is associated with a lower risk of adverse maternal outcomes when compared to the pre-Delta period.^16^ Considering the differential impact of COVID-19 variants, no study has considered the impact of all of the variants of concern thus far. In addition, currently there is no systematic study such as a meta-analysis across geographical regions with confirmed SARS-CoV-2 infections during pregnancy to assess current research and derive conclusions. With an overall aim of expanding the currently limited understanding of the impact of COVID-19 on pregnant mothers and their neonates, the main objective of this study was to conduct an individual patient data meta-analysis with collaborators from various countries to associate adverse maternal and neonatal outcomes with specific SARS-CoV-2 variants of concern.

## Methods

Our study is a secondary analysis of the data from the perinatal COVID-19 sequential prospective meta-analysis study by Smith et al^17^ (protocol was registered with PROSPERO (ID: 188955; protocol # CRD42020188955)) and was determined to be exempt from IRB review at The George Washington University. Approval from each study site was granted as follows:

Martinez-Portilla, 2021 (Mexico): Study protocol was approved by the General Hospital of Mexico (Dr Eduardo Liceaga), Mexico City, Mexico, under ethics committee number CE/23020.

Bracero, Valencia, Delgado-Lopez, 2021 (Puerto Rico): Research was conducted through the COVID-19 surveillance system under the guidance of the Puerto Rico Department of Health; no IRB review needed.

Favre, Panchaud, 2022 (Switzerland, France): Study was approved by both the Swiss Ethical Board (CER-VD-2020-00548) and the local ethics boards at each participating center. French data was registered with the French National Data Protection Commission (CNIL - authorization 2217464).

Poon, 2021 (Hong Kong, China): Study was approved by the Joint Chinese University of Hong Kong-New Territories East Cluster Clinical Research Ethics Committee. Ethical approval was granted (CREC Ref. No. 2020.210).

Bevilacqua, Laurita Longo, 2020 (Rome, Italy): Study was approved by the Universita Cattolica del Sacro Cuore (Approval number: No PROT.APROV.IST DIPUSVSP-24.02-217).

Akelo, Tippett Barr, 2021 (Kenya): Study was approved by the Kenya Medical Research Institute (KEMRI) (SERU # 4166). Study was also cleared by the CDC in Kenya (CDC Project ID # 0900f3eb81c36d6c).

Gil, Fernandez Buhigas, 2021 (Madrid, Spain): Study was approved by the Comité de Ética de la Investigación con Medicamentos de los Hospitales Universitarios de Torrevieja y Elche-Vinaolopó, c/ Tónico Sansano Mora 14, 03293, Elche, Alicante, affiliated with Hospital Universitario del Vinalopó (Approval number: 2020.03; Date of approval: 17 June 2020).

### Eligibility criteria

Eligible studies include registries and single- or multi-site cohort studies that recruited pregnant and recently postpartum women with confirmed or suspected COVID-19. Studies must have enrolled at least 25 women within a defined catchment area. Studies also had to have data that overlapped more than a single COVID-19 variant time period.

### Study selection

We invited principal investigators of studies of COVID-19 in pregnancy who were already participating in the prospective meta-analysis by Smith & Oakley et al.^17^

### Data extraction and IPD Integrity

Following identification of eligible studies, investigators shared individual patient data (IPD) with the technical team for review and analysis. The technical team processed data to review data quality, identify outliers, and reconstruct variables to align with harmonized definitions of outcomes as defined in our protocol. Study sites that were unable to share IPD directly submitted aggregate results after following the same IPD data analysis process set forth in the PMA.

### Outcomes

We examined 31 outcomes related to: i) COVID-19 severity (n=5); ii) maternal morbidities (n=14); iii) fetal and neonatal morbidity and mortality (n=5) and iv) adverse birth outcomes (n=8). We created two additional outcomes, maternal composite outcome and fetal composite outcome. Specific definitions of each outcome, as well as alternative definitions used in sensitivity analyses are presented in Table 2. The definitions of maternal,^18^ fetal,^19^ and neonatal death^20^ and adverse birth outcomes (low birthweight, small-for-gestational age, and preterm birth) were based on WHO case definitions.^18,19,21,22^ Individual study sites defined hospitalization, critical care, and maternal morbidity outcomes. For maternal morbidities, fetal and neonatal mortality, and all birth outcomes, we restricted to cases of COVID-19 with infection onset during pregnancy or within 7 days of pregnancy outcome, excluding postpartum cases with COVID-19 onset 8-42 days postpartum. Cases with unknown gestational age at onset were included in the analysis of pregnancy-specific outcomes and were assumed to be infections during pregnancy based on study design.

**Table 1.** Description of studies contributing to the individual patient data and aggregate data for meta-analysis.

| Author, Year | Country | Total pregnancies | Live births | Mean age (SD) | Gestational Age at Infection |  |  |  |  | Hospitalized (%) | Admitted to ICU (%) | 1st Vaccination rate (%) | Data collection timeframe |
| --- | --- | --- | --- | --- | --- | --- | --- | --- | --- | --- | --- | --- | --- |
|  |  |  |  |  | 1st Tri (%) | 2nd Tri (%) | 3rd Tri (%) | Postpartum (%) | Unknown (%) |  |  |  |  |
| Martinez-Portilla, 2021 | Mexico | 11,031 | n/a | 28.5 (6.0) | n/a | n/a | n/a | n/a | 100% | 20% | 2% | n/a | Mar2020-Mar2021 |
| Bracero, Valencia, Delgado-Lopez, 2021 | Puerto Rico, USA | 2,022 | 1758 | 26.6 (5.6) | 9% | 15% | 30% | n/a | 45% | n/a | 1% | n/a | Mar2020-May2022 |
| Favre, Panchaud, 2022 | Switzerland* | 1,231 | 1,124 | 31.7 (5.0) | 15.3% | 32.4% | 50.6% | n/a | 1.8% | n/a | 3% | 0% | Mar 2020-Aug 2022 |
| Favre, Panchaud, 2022 | France* | 855 | 844 | 31.4 (5.0) | 11.6% | 35.6% | 52.6% | n/a | 0.2% | n/a | 5% | 0% | Mar 2020-Aug 2022 |
| Poon, 2021 | Hong Kong, China | 121 | 120 | 32.7 (5.5) | 3% | 9% | 85% | 1% | 2% | 93% | 2% | 25% | Mar2020-Mar2022 |
| Bevilacqua, Laurita Longo, 2020 | Rome, Italy | 427 | 413 | 33.0 (5.6) | 3% | 3% | 92% | 1% | 1% | 8% | 2% | 16% | Feb2020-Mar2022 |
| Akelo, Tippet Barr, 2021 | Kenya | 125 | 91 | 26.4 (5.2) | 1% | 12% | 30% | 29% | 28% | 9% | 0% | n/a | Aug2020-May2021 |
| Gil, Fernandez Buhigas, 2021 | Madrid, Spain | 517 | 402 | 32.8 (5.4) | 25% | 39% | 35% | n/a | 1% | 3% | n/a | 37% | Jan2020-Mar2022 |

\*Submitted aggregate results

**Table 2.**
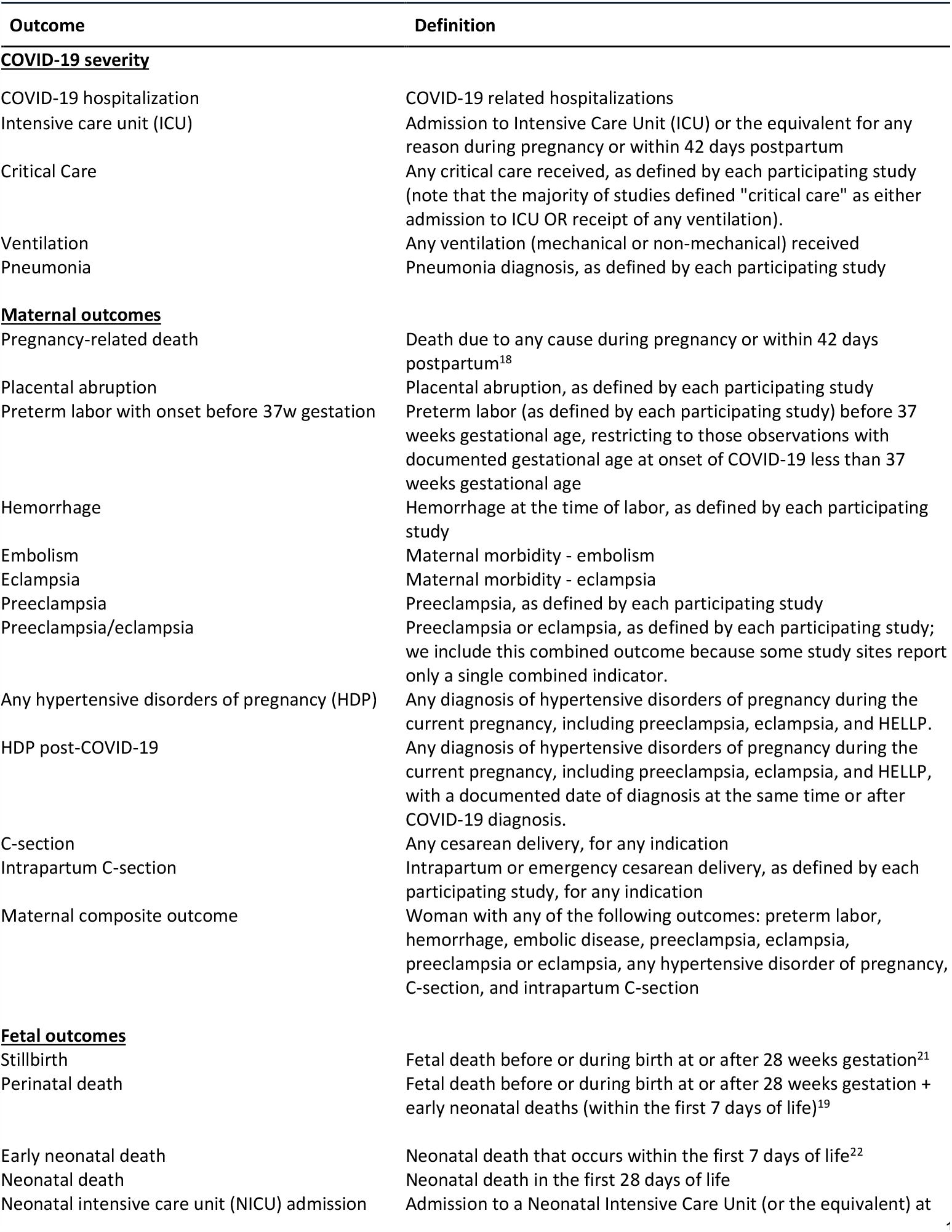
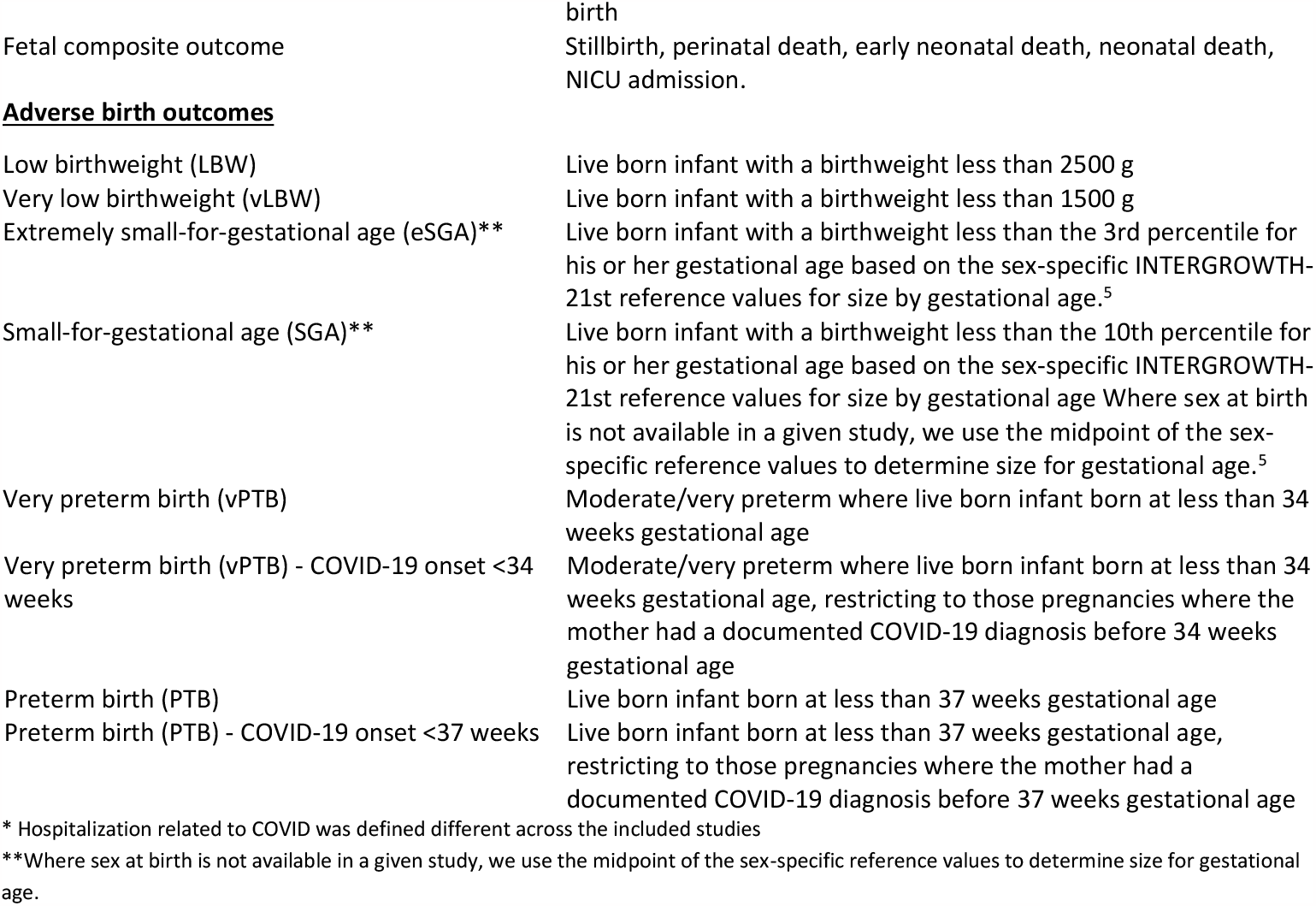
Outcome definitions^17^.

#### Exposure

SARS-CoV-2 strains that have been identified as variants of concern (VOC) by the WHO23 were analyzed using publicly available strain frequency data from Nextstrain.org. VOCs identified and analyzed in this study are Pre-alpha, Alpha, Beta, Delta, Epsilon, Eta, and Omicron. For each month, the dominant strain was defined to be the strain that was ≥50% prevalent at the country level, consistent with the CDC definition of strain dominance.24 If no strain reached this threshold, the dominant strain was labeled as “Mixed.” We also conducted sensitivity analyses with two additional thresholds (≥60% prevalence and ≥70% prevalence) in categorizing a strain as the dominant strain.

Nextstrain is an open-source project that compiles and publishes frequency data on VOCs by month and country.^25^ Genetic data are submitted by researchers to Nextstrain, where a COVID-19 variant lineage is tracked for each available country on a weekly basis. Strain frequency data downloaded as JSON was converted to CSV using Python version 3.9.13 and analyzed in R version 4.2.0. Nextstrain data was last retrieved October 2022.

Two countries had short stretches of missing data in the publicly available Nextstrain (https://nextstrain.org) dataset; in our study, Madrid, Spain contributed data through March 2022 but variant data from Nextstrain was only available until January 2022; hence, study data past January 2022 was not included in the analysis. Hong Kong SAR (China) Nextstrain data was missing for August 2021 and the data collected during that month was dropped in our analysis.

### Statistical Analysis

Individual patient data from six countries (Rome (Italy), Mexico, Puerto Rico (USA), Madrid (Spain), Kenya, Hong Kong SAR (China)) and aggregate data from two countries (Switzerland and France) was merged with the frequency data from Nextstrain.org based on country, month, and year.

### Data synthesis

We applied a 2-stage IPD meta-analytic framework, using the R Meta package (version 5.5.0).^26^ Using this method, we first calculated study-specific estimates for each outcome, for each possible comparison of one dominant strain versus another dominant strain (all combinations of Pre-alpha, Beta, Delta, Epsilon, Eta, Omicron, and Mixed) in a pairwise fashion. This generated proportions and relative risks (RR) with 95% confidence intervals (CI) for each participating study, outcome, and strain comparison. We then generated pooled relative risks, with 95% CI for each dominant variant and outcome pair when there were one or more studies with available data (individual study data not shown here). The R Meta package uses the generic inverse variance method, where larger weights are given to bigger studies than smaller studies, which have larger standard errors,^27^ to generate pooled relative risks and to assess heterogeneity across studies using the I^2^ statistic. We present unadjusted estimates because the goal of this study was to present descriptive epidemiological data among pregnant women with COVID-19 and their infants, rather than to examine a causal relationship.

We excluded studies that had zero total events in a particular analysis. In the case of zero events within a variant subgroup, we added a continuity correction of 0.5 to each cell in the contingency table when calculating pooled relative risks. All meta-analyses were conducted in R version 4.2.0.^28^ and the code can be found on GitHub: https://github.com/SmithLabGWSPH/COVID-19-Variant-PMA.

## Results

### Study selection

We included data from studies across eight countries (Italy, Mexico, Puerto Rico, Spain, Kenya, China-Hong Kong, Switzerland, and France) with confirmed SARS-CoV-2 cases in pregnancy or postpartum. Table 1 shows a description of studies contributing to the individual patient data meta-analysis. Our analysis included data from any study that met the eligibility criteria and were able to share data by November 2022 (Figure 1). Favre, Panchaud, 2022 contributed data from two countries (Switzerland and France) and data was therefore analyzed as two separate studies.

**Figure 1.**
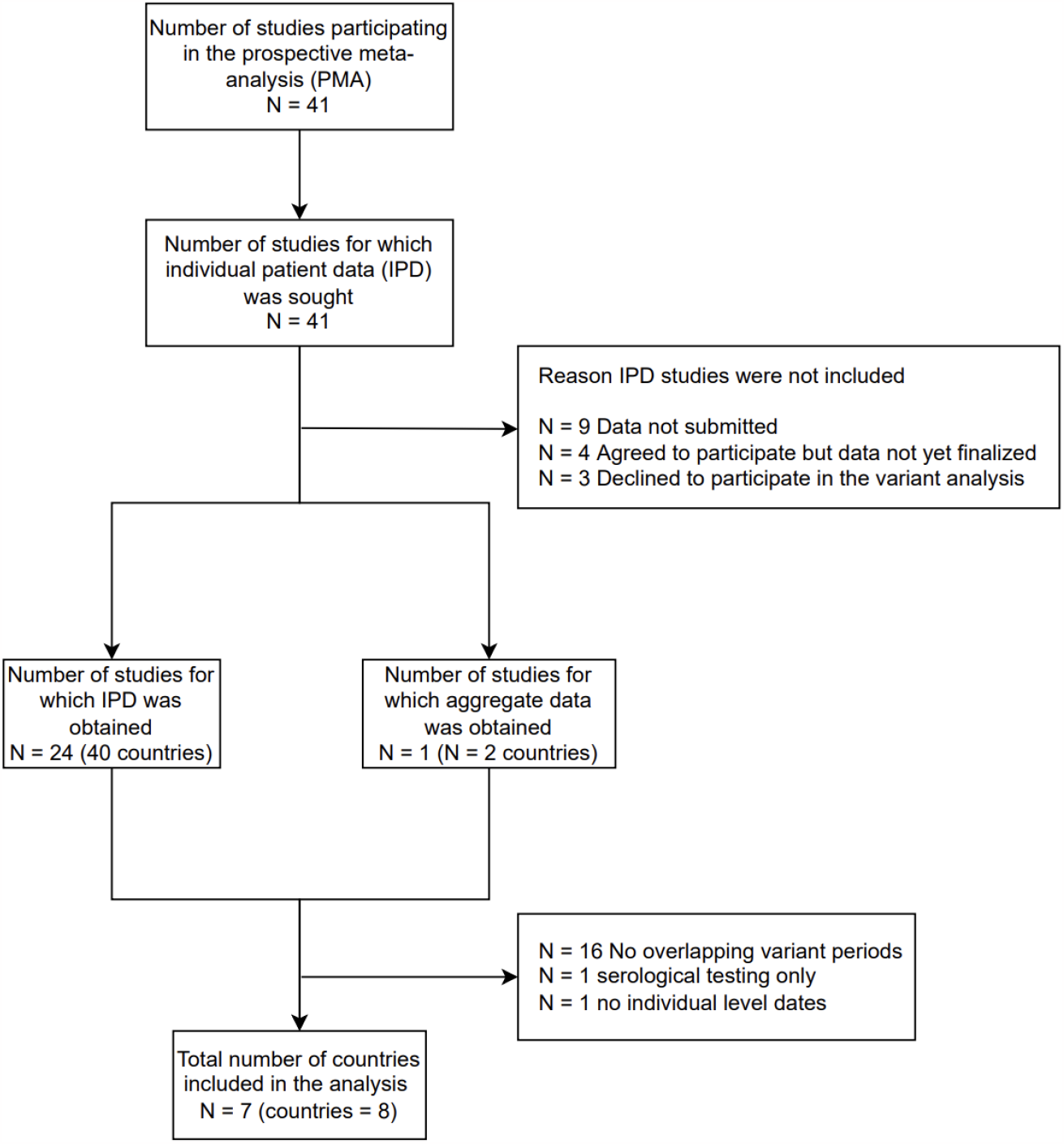
PRISMA Flow Chart for COVID-19 Variant Studies.

### Study characteristics

Data collection for each study spanning the variant time period is shown in Figure 2. Across the eight studies, a total of 16,403 pregnancies with confirmed SARS-CoV-2 infection were observed. More than 11,000 cases were contributed by the Mexico National Registry (Martinez-Portilla, 2021) and the rest were contributed by the other 7 countries (Table 1). Mean age ranged from 26.4±5.2 years in Kenya to 33.0±5.6 years in Rome, Italy. The majority of the studies recruited pregnant women during the third trimester, with four studies recruiting participants during the postpartum period as well. Rate of first vaccination was documented in three of the eight studies, with the highest coverage (37%) in Madrid, Spain. Data collection time frame varied from nine months (August 2020 - May 2021 in Kenya) during the pandemic to two years and three months (Jan 2020 to March 2022 in Madrid, Spain).

**Figure 2.**
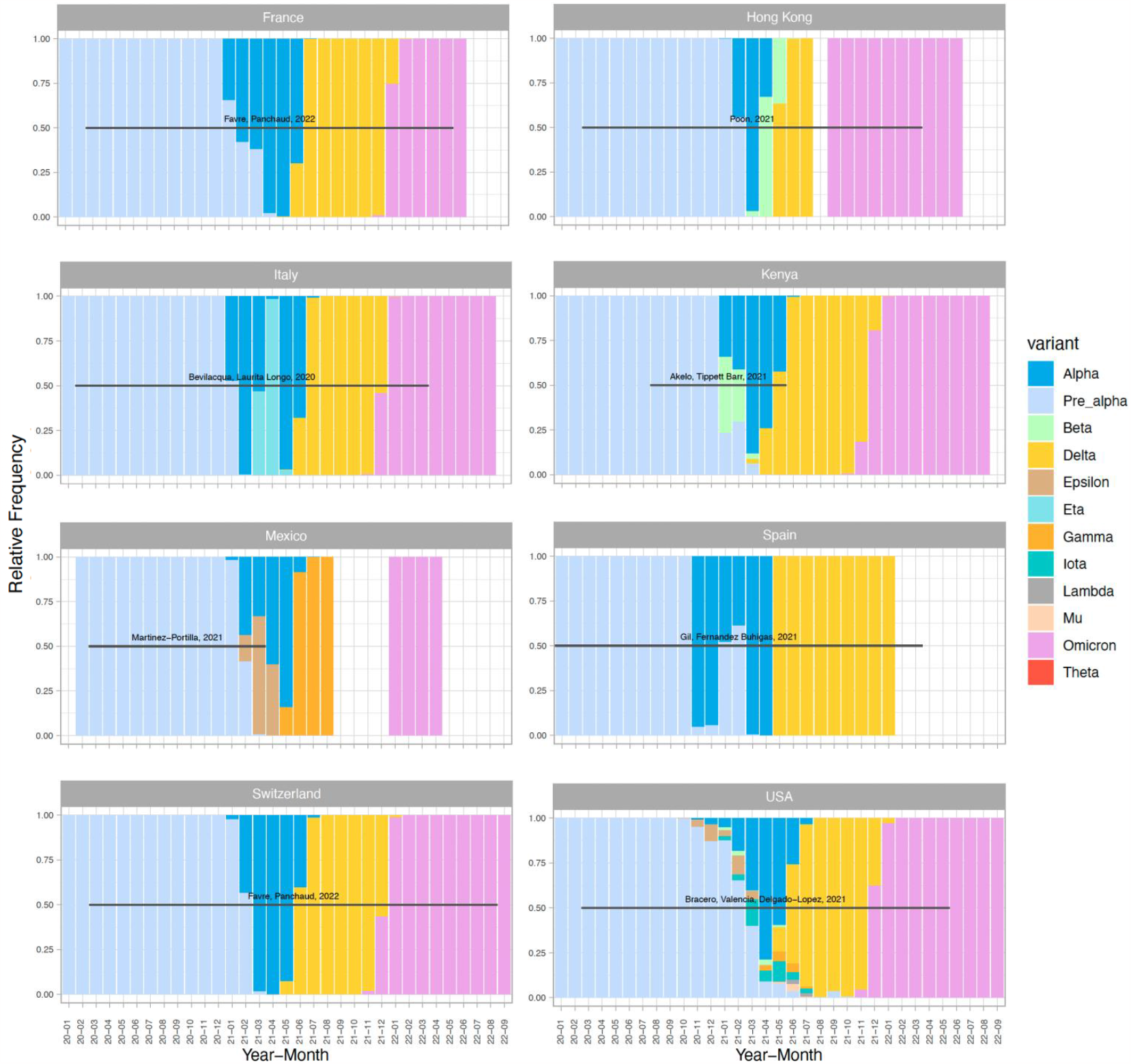
Prevalent strain frequency (nextstrain.org) with overlapping study data during months of the pandemic (Jan 2020 - Sep 2022)

### Study estimates

Table 3 (A-D) and Figure 3 (A-F) show pooled relative risks with 95% confidence intervals for each pair of COVID-19 variants for each outcome where data was available. Table 3A shows that the Delta wave had a higher risk of all adverse COVID-19 severity outcomes compared to the Omicron wave. Risk of hospitalization was [RR 4.02 (95% CI 1.10, 14.69), n=1 study] during the Delta wave compared to the Omicron wave. Additionally, during the Delta wave, risk of ICU admissions was [RR 2.59 (95% CI 1.26, 5.30), n=3 studies], risk of critical care admission was [RR 2.52 (95% CI 1.25, 5.08), n=3 studies], risk of needing ventilation was [RR 3.96 (95% CI 1.47, 10.71), n=3 studies] and risk of pneumonia was [RR 6.73 (95% CI 2.17, 20.90), n=3 studies], compared to the Omicron wave. Additionally, risks of ICU admission, critical care and requiring ventilation during the Delta wave were higher compared to the Pre-alpha wave ([RR 2.15 (95% CI 1.04, 4.47), n=3 studies], [RR 1.96 (95% CI 1.06, 3.62), n=4 studies], and [RR 2.69 (95% CI 1.14, 6.33), n = 4], respectively). There was one study showing increased risk for hospitalization during the Epsilon wave compared to the Pre-alpha wave [RR 1.57 (95% CI 1.15, 2.14), n=1 study] and four studies showing an increased risk for ICU admissions [RR 2.83 (95% CI 1.10, 7.26), n = 4 studies] and ventilation [RR 3.20 (95% 1.22, 8.44), n = 4 studies] during the Alpha wave compared to the Omicron wave. Lastly, compared to the Omicron wave, Pre-Alpha period infections showed increased risk for pneumonia [RR 3.14 (95% CI 1.06, 9.26), n=3 studies].

**Table 3.**
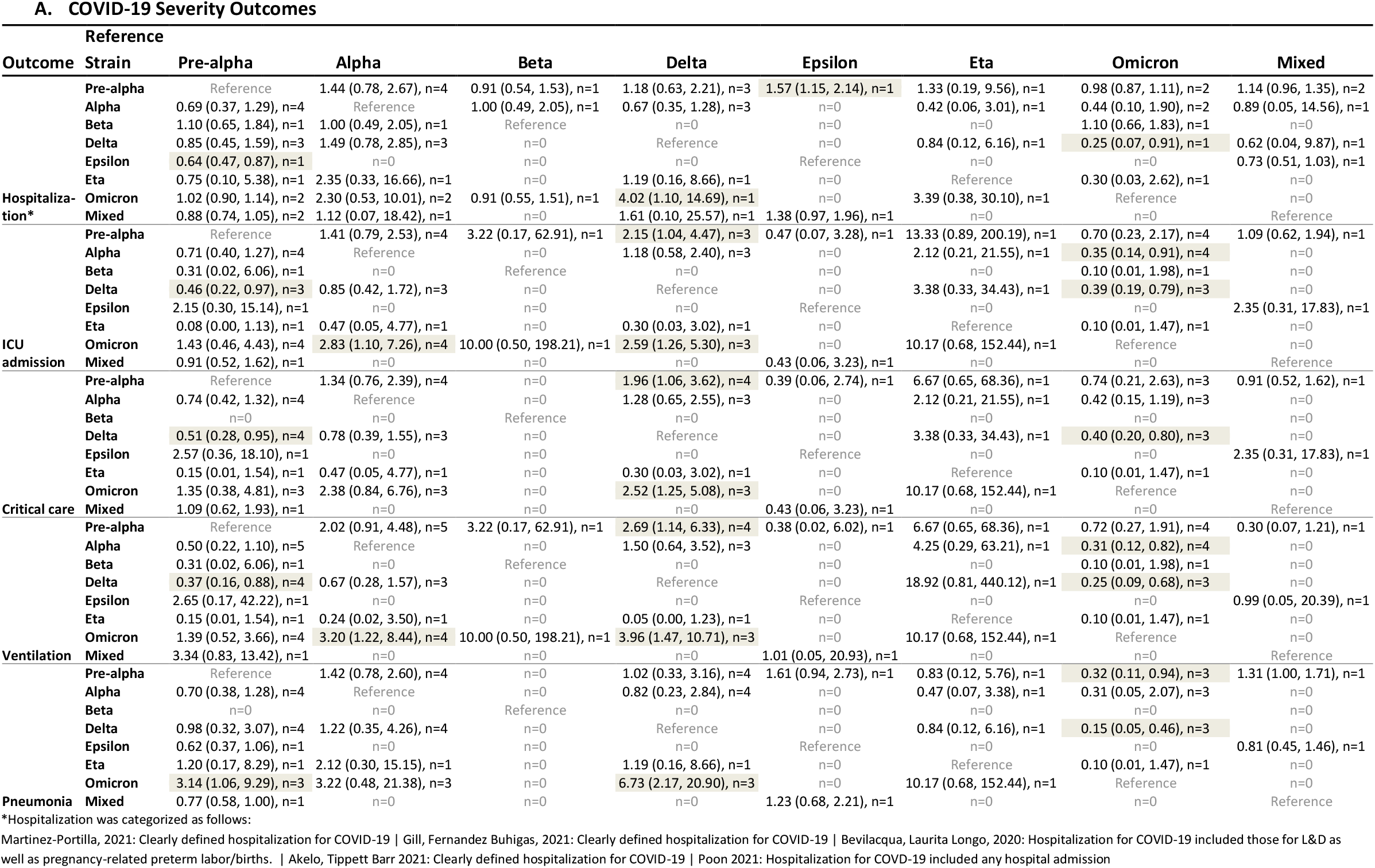
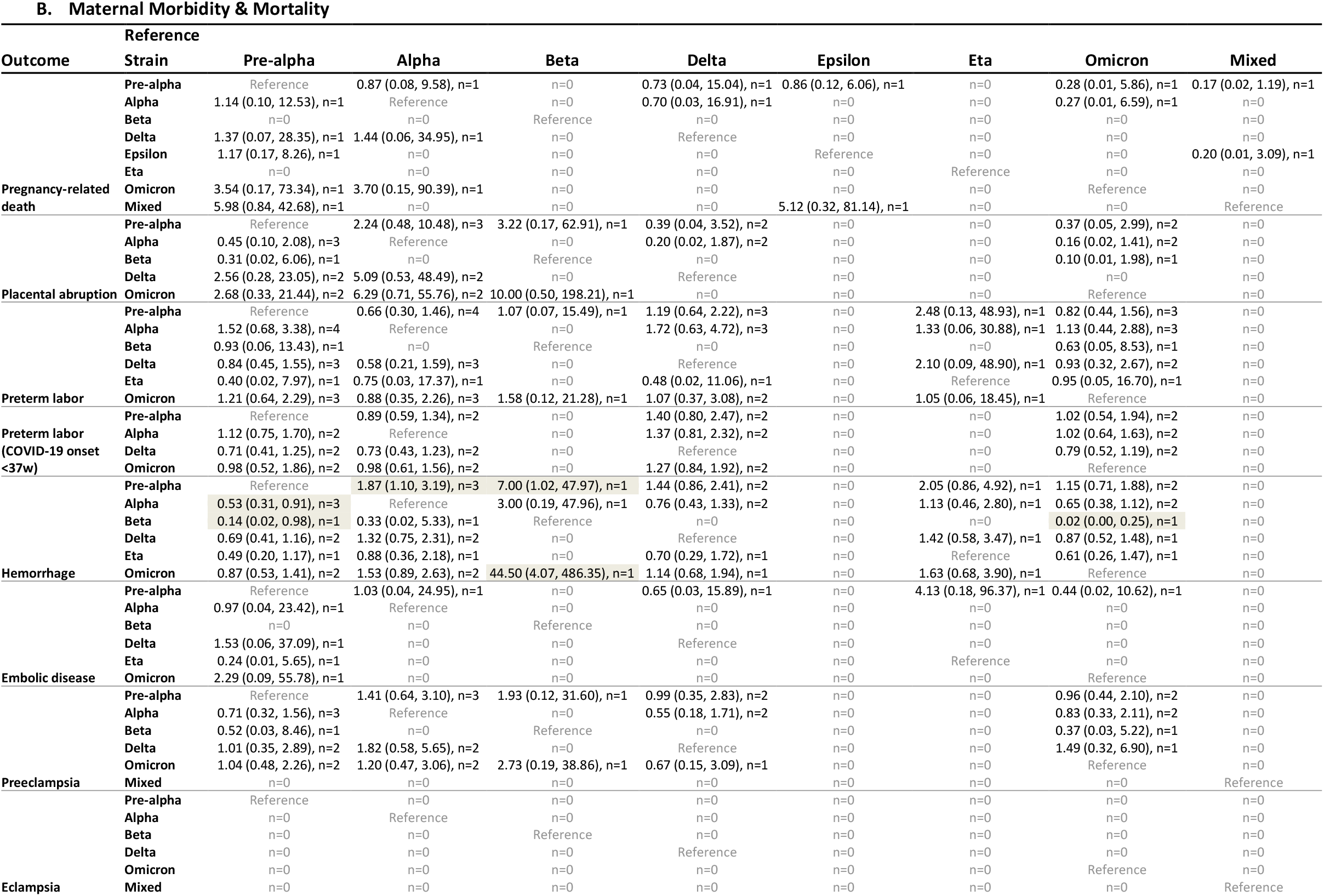
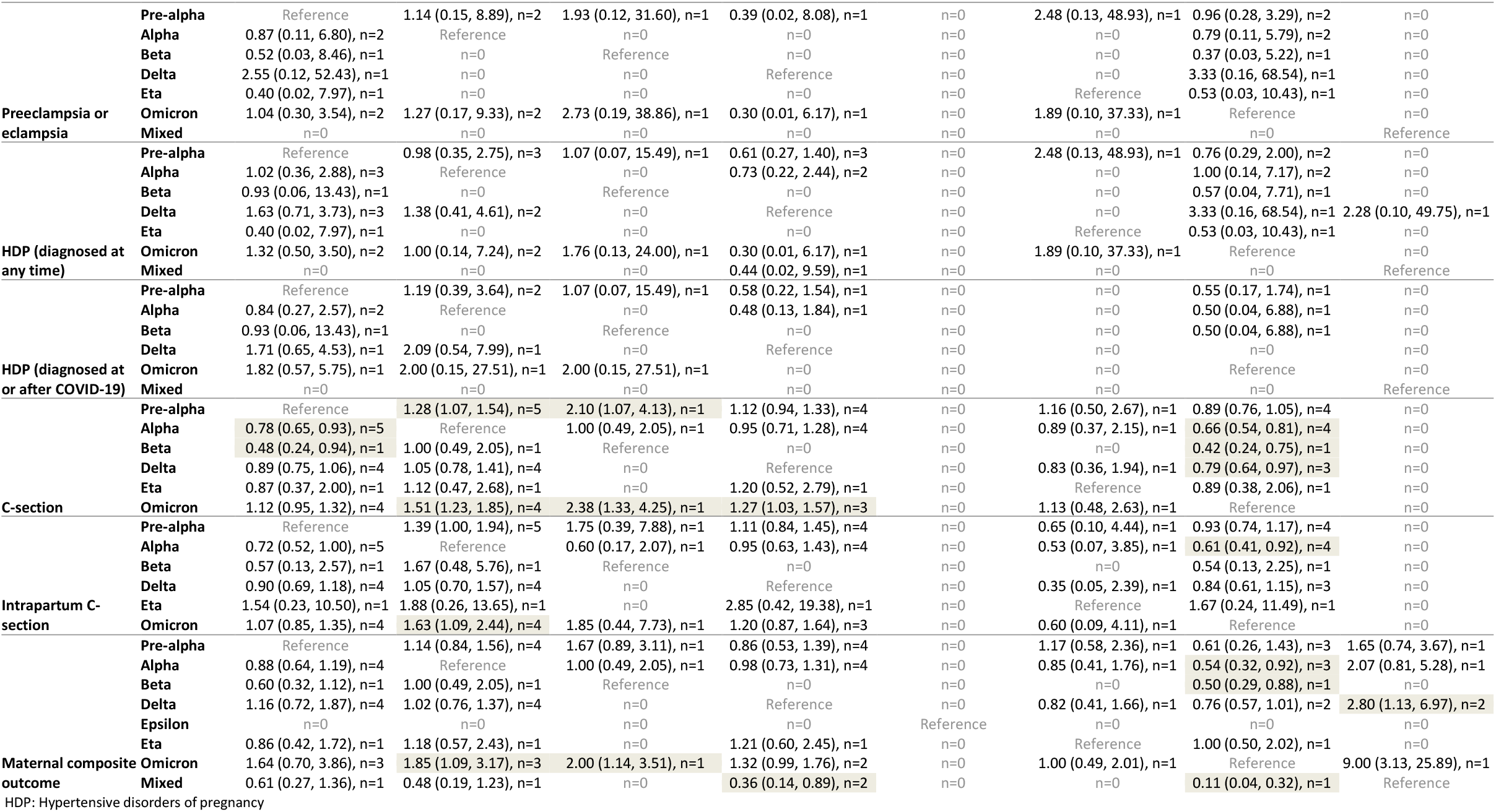
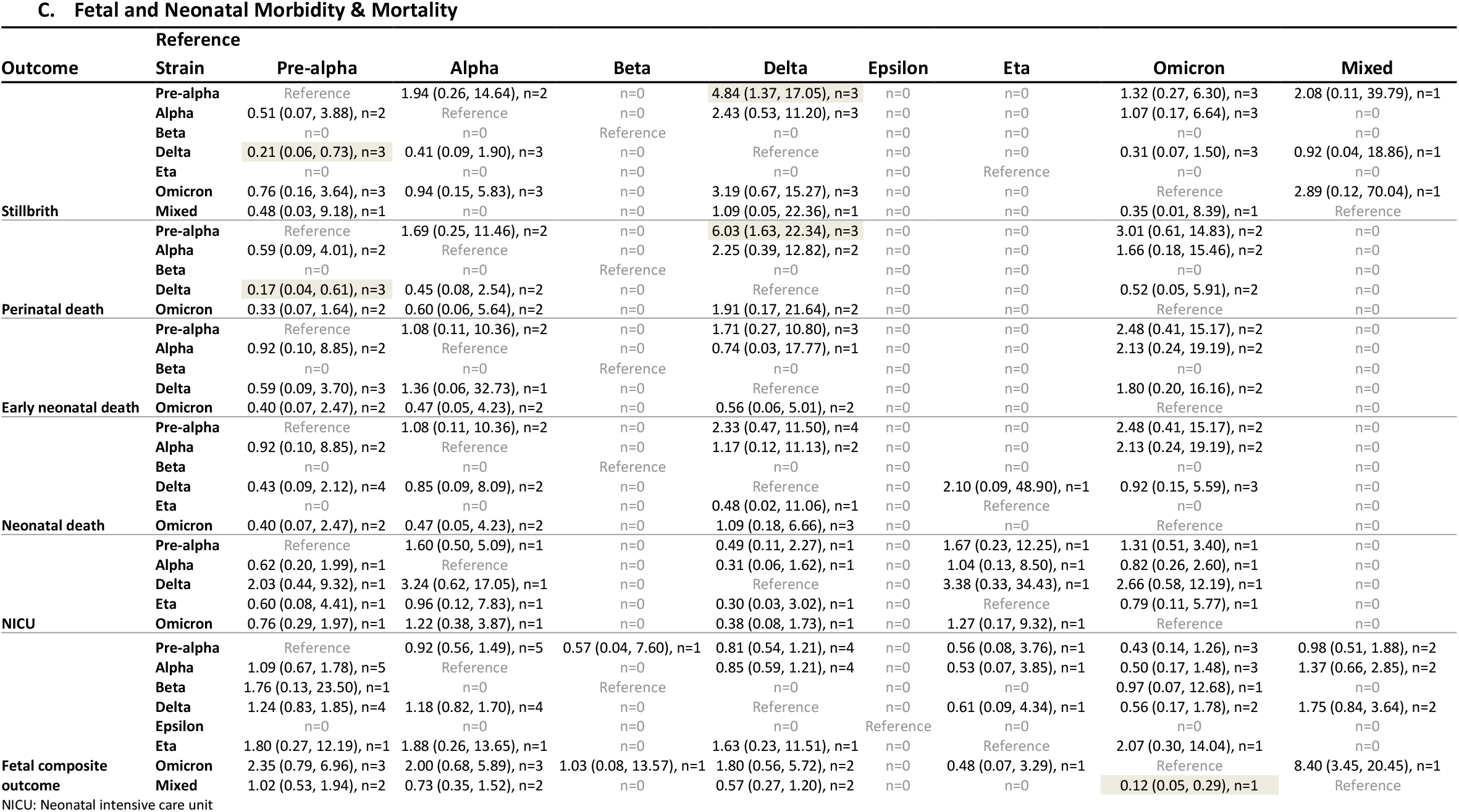
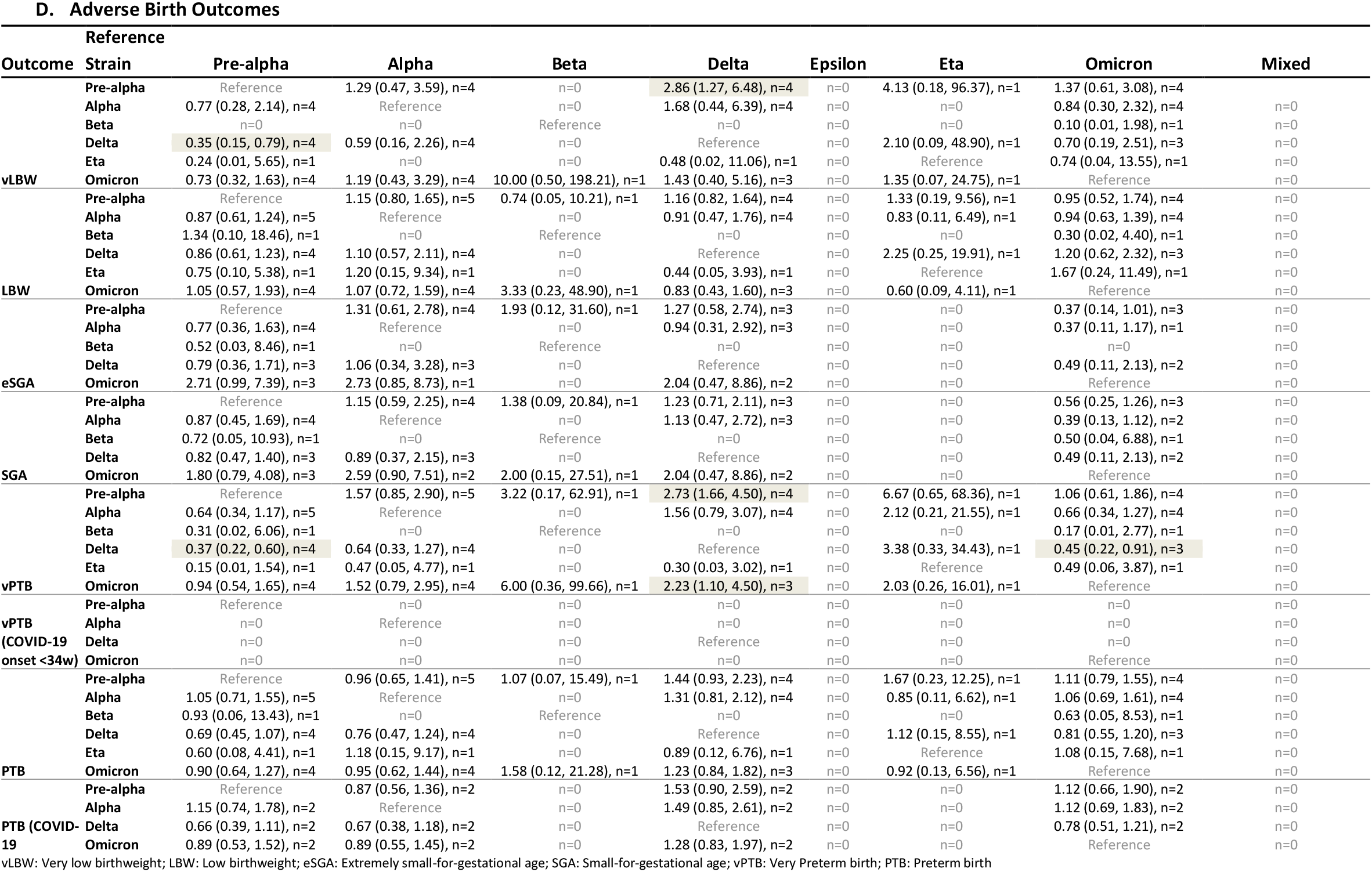
Random effect relative risk (RR) estimates with 95% confidence intervals (CI) and number of studies included in each analysis.

**Figure 3.**
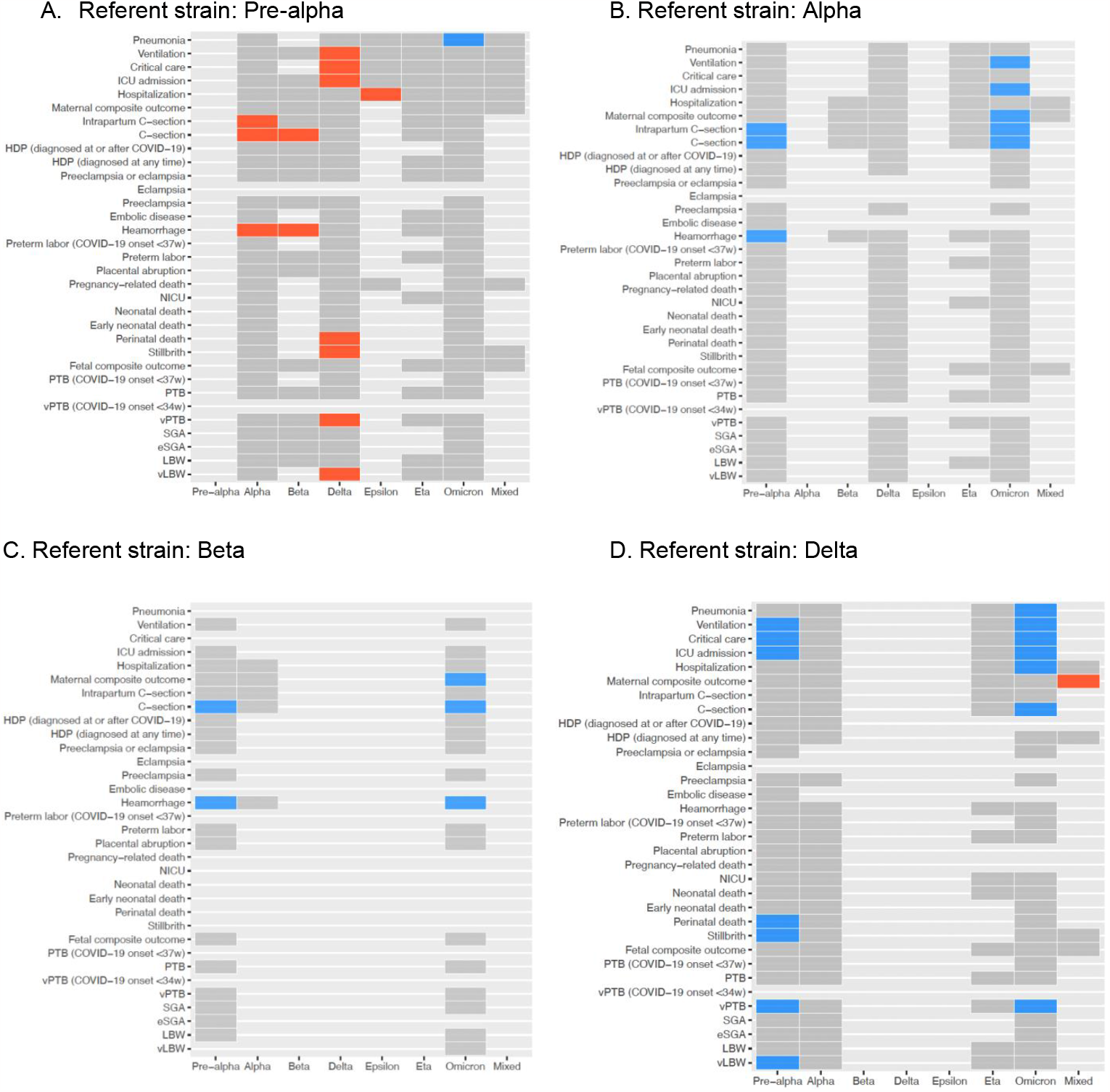
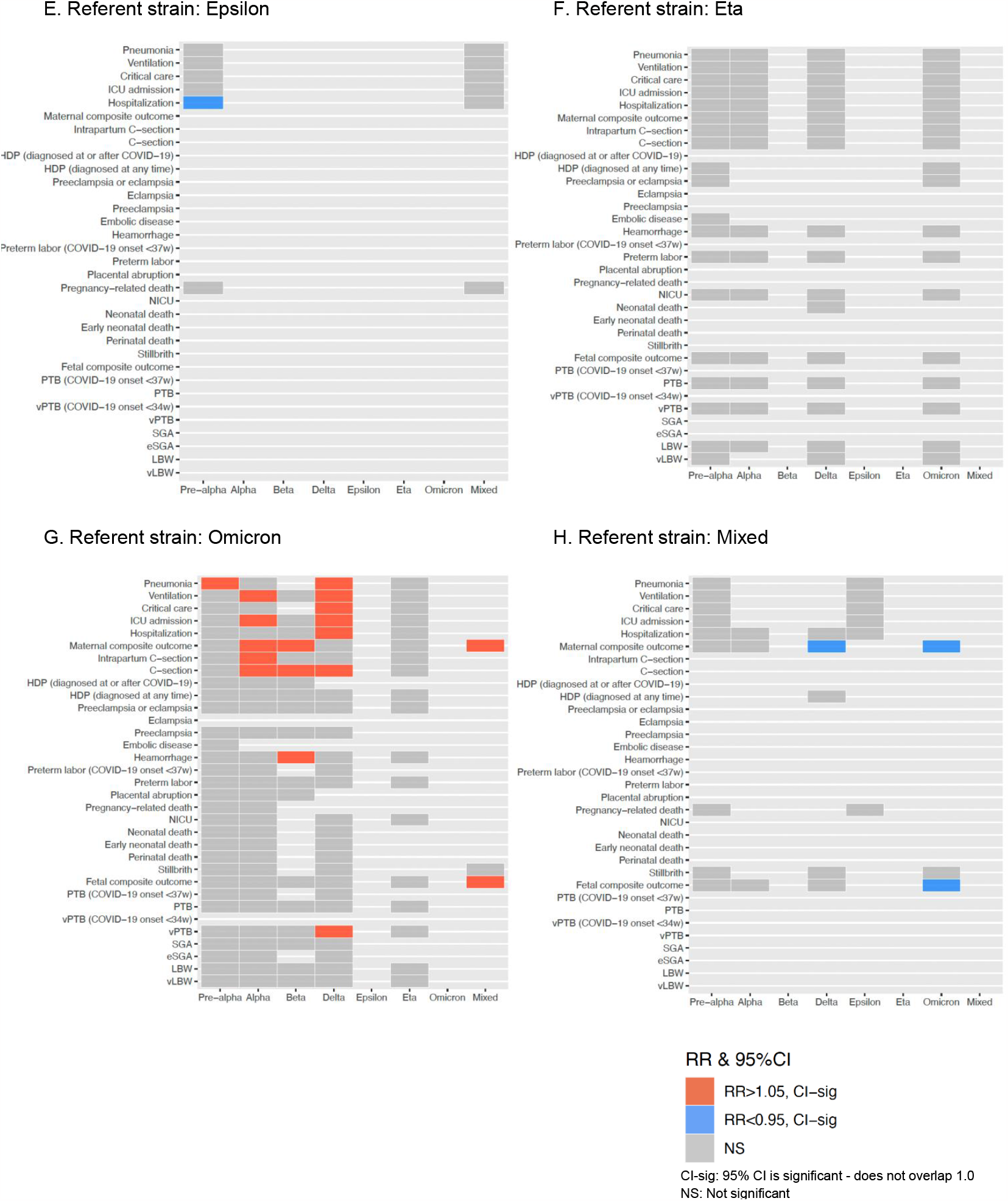
Tile map showing significant associations between referent strain and test strains (Dominant strain≥50%)

Our results for maternal morbidity and mortality (Table 3B) indicators, showed that no significant increased risk during any of the COVID-19 variant waves except hemorrhage, C-section, and intrapartum C-section and maternal composite outcome. During the Alpha wave, pregnant women were at an increased risk for hemorrhage [RR 1.87 (95% CI 1.10, 3.19), n=3 studies] and C-section [RR 1.28 (95% CI 1.07, 1.54), n=5 studies] compared to the Pre-alpha wave. Risk of any C-section and risk of intrapartum C-section was also increased during the Alpha wave compared to the Omicron wave [RR 1.51 (95% CI 1.23, 1.85) n= 4 studies and RR 1.63 (95% CI 1.09, 2.44), n = 4 studies, respectively]. Further, the risk of maternal composite outcome was elevated during the Alpha wave compared to Omicron [RR 1.85 (95% CI 1.09, 3.17), n = 3 studies]. Risk of hemorrhage [RR 7.00 (95% CI 1.02, 47.97), n=1 study] and C-section [RR 2.10 (95% CI 1.07, 4.13), n=1 study] was also increased during the Beta wave, each in just one study, compared to the Pre-alpha wave.

Additionally, risk of hemorrhage, C-Section, and maternal composite outcome, was increased during the Beta wave compared to the Omicron wave ([RR 44.50 (95% CI 4.07, 486.35) n=1 study], [RR 2.38 (95% 1.33, 4.25) n=1 study], [RR 2.00 (95% CI 1.09, 3.17), n=1 study], respectively). The Delta wave showed an increased risk for C-sections [RR 1.27 (95%CI 1.03, 1.57), n = 3 studies] when compared to the Omicron wave. Mixed time periods, when no single variant comprised more than 50% of the caseload, showed significant increased risks for maternal composite outcome, compared to the period of Delta predominance [RR 2.80 (95% CI 1.13, 6.97), n=2 studies] and compared to the period of Omicron predominance [RR 9.00 (95% CI 3.13, 25.89), n=1 study].

Risk of fetal and neonatal morbidity and mortality (Table 3C) did not show significant increases in risks during any of the COVID-19 waves except stillbirth and perinatal death during the Delta wave when compared to the Pre-alpha wave ([RR 4.84 (95% CI 1.37, 17.05), n=3 studies and [RR 6.03 (95%CI 1.63, 22.34), n=3 studies], respectively). Mixed variant time periods showed increased risk for fetal composite outcome when compared to the Omicron wave in a single study [RR 8.40 (95% CI 3.45, 20.45), n=1 study].

Risks of adverse birth outcomes (Table 3D) including very low birthweight and very preterm birth were increased during the Delta wave. Risk of very low birthweight [RR 2.86 (95% CI 1.27, 6.48), n=4 studies] and very preterm birth [RR 2.73 (95%CI 1.66, 4.50), n=4 studies] was higher in the Delta wave when compared to the Pre-alpha wave. Risk of very preterm birth [RR 2.23 (95% CI 1.10, 4.50), n=3 studies] was also increased in the Delta wave when compared to the Omicron wave.

Sensitivity analyses using dominant variant thresholds of ≥60% and ≥70% did not change results significantly (Supplementary Tables S1 A-D and S2 A-D).

## Discussion

Our individual-patient meta-analysis across eight countries shows that the Delta variant was significantly worse for COVID-19 severity outcomes among pregnant women and was associated with a greater incidence of perinatal death, stillbirth, very preterm birth and very low birthweight compared to the Pre-alpha and Omicron strains. Alpha strain was also worse for some COVID-19 severity outcomes as well as for a few maternal outcomes compared to the Omicron variant. Studies show that the COVID-19 pandemic has not only adversely impacted the general population, but it has also been detrimental to pregnant women and their infants.^29,30^ Some variants of the wild-type (Pre-alpha) strain have been worse than others^31,32^; research shows that the Delta variant was much more transmissible, contagious, and efficient at infection^31^ since it was first reported in India December 2020.^33^ During periods of Delta strain predominance, COVID-19 severity outcomes were more severe both among pregnant women and among non-pregnant individuals^34^, compared to the other COVID-19 waves. Our results are in line with several recent studies on COVID-19 in pregnancy showing that Delta predominance was associated with increased COVID-19 severity compared to Omicron predominance.^14,34–36^ Consistent with previous studies, our study also shows that the risk of stillbirth increased during the Delta wave compared to its predecessors (in our analysis, Pre-alpha).^11,37^ Our meta-analysis is first to compare all variants of concern with each other during pregnancy, and to do so utilizing studies from diverse geographic regions.

As a meta-analysis, our study provides more precise estimates than an individual study might be able to provide.

### Strengths and limitations

Our analysis had several notable strengths. First, IPD meta-analyses are considered the gold-standard method for generating aggregate estimates. Our outcome data from each country was standardized and harmonized based on universal definitions. We defined strict study inclusion criteria and only included studies that had confirmed SARS-CoV-2 infection during pregnancy. Another notable strength of our study is that we used a publicly available variant database for each country that uses a standardized methodology for compiling variant prevalence and is continually updated. This database not only serves as a surveillance tool but also provides a level of granularity that matches differences in health outcome patterns.

Sequencing data shared by hundreds of laboratories across the world, deposited in NCBI^38^, GISAID^39^, ViPR^40^, certain GitHub repositories and other public sources, is curated and processed in such a way that is agnostic to the source data and can therefore be used for generalized inferences.^25^ To our knowledge, this is the first systematic analysis to assess the impact of all known variants of concern on maternal and fetal outcomes during pregnancy. In the literature, studies have only compared data in the Delta wave to the Omicron wave and/or to what is considered as the Pre-delta period.

Despite many strengths, there are limitations in our study. Studies contributing to the IPD meta-analysis recruited participants using varying methods, including hospital-based surveillance and screening during antenatal care, which could bias our findings regarding COVID-related hospitalizations. Although several studies are included, the majority of data on COVID-19 severity outcomes (hospitalization, ICU admission, critical care, ventilation, and pneumonia) was comprised of data from the Mexican National Registry, which did not collect data on maternal morbidity and mortality, fetal and neonatal outcomes or adverse birth outcomes.

Data on vaccination status during pregnancy is incomplete in this dataset. Future studies are needed to assess the impact of different variants on pregnancy outcomes among those who were vaccinated compared to those who were not. Data on SARS-CoV-2 infections prior to becoming pregnant or vaccination are not available for these study populations; moreover, this data would be challenging to collect without underestimating given that some infections are asymptomatic. A reinfection during pregnancy or after vaccination would imply a change in the activation or response by the immune system. A differential activation can impact the true pathogenicity measurement and may lead to an underestimation of the real risk, especially for the Omicron and Delta variants that came later in the pandemic. Reinfections during the Omicron wave increased by 15%^41^ and data shows that the Omicron variant had the ability to evade the immune system, although it still had similar protection against hospitalizations or death due to reinfection.^42^

Our study utilizes country-level estimates to assign exposures to individual-level data; hence, further studies are needed to validate our results. However, any exposure measurement error would be nondifferential because the same method was used for all variants of concern. Using person-specific genomic data would provide the best estimates, rather than country-level variant prevalence.

Lastly, data on adverse outcomes during each variant period were especially sparse; not only are adverse outcomes generally rare, but the adverse outcomes present in the data are further subdivided among several variants for the purpose of comparison. In order to increase sample size and improve the power of the pooled estimates, efforts should be made to expand the meta-analysis with additional, existing studies on COVID-19 in pregnancy.

### Conclusion

Understanding how different variants of pathogens impact the health of the population is important to mitigating public health emergencies, such as the current pandemic. Genomic surveillance and analysis help monitor diseases and tailor interventions and recommendations for the general public. In the field of public health, there are numerous examples of respiratory illnesses that rapidly evolve to the detriment of vulnerable groups such as the elderly, children under-five, and pregnant women. The influenza A virus pandemic (pH1N1)^43^ of 2009 was a recent example with a higher prevalence among young adults that gradually evolved to higher attack rates among older adults^44^; data shows that pH1N1 adversely impacted pregnancy outcomes as well, especially birthweight in infants.^45^ Additionally, the current coronavirus pandemic exposed gaps in national health systems and responses. National health agencies and researchers must continue to collaborate to monitor and analyze multi-country variant data in order to address these gaps and mitigate adverse health consequences amidst ongoing and future outbreaks.

## Data Availability

All data produced in the present study are available upon reasonable request to the authors

## Footnotes

1a We refer to all pregnant people as pregnant women throughout this study.

## References

1. Harrison, A. G., Lin, T. & Wang, P. Mechanisms of SARS-CoV-2 Transmission and Pathogenesis. Trends Immunol. 41, 1100–1115 (2020).

2. Marian Knight, Rema Ramakrishnan, Kathryn Bunch, Nicola Vousden, Jennifer J Kurinczuk, Sarah Dunn, Lisa Norman, Aisling Barry, Ewen Harrison, Annemarie Docherty, Calum Semple. Females in hospital with SARS-CoV-2 infection, the association with pregnancy and pregnancy outcomes A UKOSS/ISARIC/CO-CIN investigation.

3. Samara, A., Khalil, A., O’Brien, P. & Herlenius, E. The effect of the delta SARS-CoV-2 variant on maternal infection and pregnancy. iScience 25, 104295 (2022).

4. Vousden, N., Ramakrishnan, R., Bunch, K., Morris, E., Simpson, N. A. B., Gale, C., O’Brien, P., Quigley, M., Brocklehurst, P., Kurinczuk, J. J. & Knight, M. Severity of maternal infection and perinatal outcomes during periods of SARS-CoV-2 wildtype, alpha, and delta variant dominance in the UK: prospective cohort study. BMJ Medicine 1, (2022).

5. Villar, J., Ariff, S., Gunier, R. B., Thiruvengadam, R., Rauch, S., Kholin, A., Roggero, P., Prefumo, F., do Vale, M. S., Cardona-Perez, J. A., Maiz, N., Cetin, I., Savasi, V., Deruelle, P., Easter, S. R., Sichitiu, J., Soto Conti, C. P., Ernawati, E., Mhatre, M., Teji, J. S., Liu, B., Capelli, C., Oberto, M., Salazar, L., Gravett, M. G., Cavoretto, P. I., Nachinab, V. B., Galadanci, H., Oros, D., Ayede, A. I., Sentilhes, L., Bako, B., Savorani, M., Cena, H., García-May, P. K., Etuk, S., Casale, R., Abd-Elsalam, S., Ikenoue, S., Aminu, M. B., Vecciarelli, C., Duro, E. A., Usman, M. A., John-Akinola, Y., Nieto, R., Ferrazi, E., Bhutta, Z. A., Langer, A., Kennedy, S. H. & Papageorghiou, A. T. aternal and Neonatal Morbidity and Mortality Among Pregnant Women With and Without COVID-19 Infection: The INTERCOVID Multinational Cohort Study. JAMA Pediatr. 175, 817–826 (2021).

6. Allotey, J., Stallings, E., Bonet, M., Yap, M., Chatterjee, S., Kew, T., Debenham, L., Llavall, A. C., Dixit, A., Zhou, D., Balaji, R., Lee, S. I., Qiu, X., Yuan, M., Coomar, D., Sheikh, J., Lawson, H., Ansari, K., van Wely, M., van Leeuwen, E., Kostova, E., Kunst, H., Khalil, A., Tiberi, S., Brizuela, V., Broutet, N., Kara, E., Kim, C. R., Thorson, A., Oladapo, O. T., Mofenson, L., Zamora, J., Thangaratinam, S. & for PregCOV-19 Living Systematic Review Consortium. Clinical manifestations, risk factors, and maternal and perinatal outcomes of coronavirus disease 2019 in pregnancy: living systematic review and meta-analysis. BMJ 370, m3320 (2020).

7. Smith, E. R., Oakley, E., Grandner, G. W., Ferguson, K., Farooq, F., Afshar, Y., Ahlberg, M., Ahmadzia, H., Akelo, V., Aldrovandi, G., Tippett Barr, B. A., Bevilacqua, E., Brandt, J. S., Broutet, N., Fernández Buhigas, I., Carrillo, J., Clifton, R., Conry, J., Cosmi, E., Crispi, F., Crovetto, F., Delgado-López, C., Divakar, H., Driscoll, A. J., Favre, G., Flaherman, V. J., Gale, C., Gil, M. M., Gottlieb, S. L., Gratacós, E., Hernandez, O., Jones, S., Kalafat, E., Khagayi, S., Knight, M., Kotloff, K., Lanzone, A., Le Doare, K., Lees, C., Litman, E., Lokken, E. M., Laurita Longo, V., Madhi, S. A., Magee, L. A., Martinez-Portilla, R. J., McClure, E. M., Metz, T. D., Miller, E. S., Money, D., Moungmaithong, S., Mullins, E., Nachega, J. B., Nunes, M. C., Onyango, D., Panchaud, A., Poon, L. C., Raiten, D., Regan, L., Rukundo, G., Sahota, D., Sakowicz, A., Sanin-Blair, J., Söderling, J., Stephansson, O., Temmerman, M., Thorson, A., Tolosa, J. E., Townson, J., Valencia-Prado, M., Visentin, S., von Dadelszen, P., Adams Waldorf, K., Whitehead, C., Yassa, M., Tielsch, J. M., Perinatal COVID PMA Study Collaborators & and Perinatal COVID PMA Study Collaborators. Adverse maternal, fetal, and newborn outcomes among pregnant women with SARS-CoV-2 infection: an individual participant data meta-analysis. BMJ Glob Health 8, (2023).

8. Salvatore, C. M., Han, J.-Y., Acker, K. P., Tiwari, P., Jin, J., Brandler, M., Cangemi, C., Gordon, L., Parow, A., DiPace, J. & DeLaMora, P. Neonatal management and outcomes during the COVID-19 pandemic: an observation cohort study. Lancet Child Adolesc Health 4, 721–727 (2020).

9. Murphy, C. A., O’Reilly, D. P., Edebiri, O., Donnelly, J. C., McCallion, N., Drew, R. J. & Ferguson, W. The Effect of COVID-19 Infection During Pregnancy; Evaluating Neonatal Outcomes and the Impact of the B.1.1.7. Variant. Pediatr. Infect. Dis. J. 40, e475–e481 (2021).

10. Knight, M., Bunch, K., Vousden, N., Morris, E., Simpson, N., Gale, C., O’Brien, P., Quigley, M., Brocklehurst, P., Kurinczuk, J. J. & UK Obstetric Surveillance System SARS-CoV-2 Infection in Pregnancy Collaborative Group. Characteristics and outcomes of pregnant women admitted to hospital with confirmed SARS-CoV-2 infection in UK: national population based cohort study. BMJ 369, m2107 (2020).

11. DeSisto, C. L., Wallace, B., Simeone, R. M., Polen, K., Ko, J. Y., Meaney-Delman, D. & Ellington, S. R. Risk for Stillbirth Among Women With and Without COVID-19 at Delivery Hospitalization -United States, March 2020-September 2021. MMWR Morb. Mortal. Wkly. Rep. 70, 1640–1645 (2021).

12. Lin, L., Liu, Y., Tang, X. & He, D. The Disease Severity and Clinical Outcomes of the SARS-CoV-2 Variants of Concern. Front Public Health 9, 775224 (2021).

13. Rosenbloom, J. I., Raghuraman, N., Carter, E. B. & Kelly, J. C. Coronavirus disease 2019 infection and hypertensive disorders of pregnancy. Am. J. Obstet. Gynecol. 224, 623–624 (2021).

14. Mamun, M. M. A. & Khan, M. R. COVID-19 Delta Variant-of-Concern: A Real Concern for Pregnant Women With Gestational Diabetes Mellitus. Front. Endocrinol. 12, 778911 (2021).

15. CDC. SARS-CoV-2 variant classifications and definitions. Centers for Disease Control and Prevention (2022). at <https://www.cdc.gov/coronavirus/2019-ncov/variants/variant-classifications.html>

16. Favre, G., Maisonneuve, E., Pomar, L., Daire, C., Poncelet, C., Quibel, T., Monod, C., Martinez de Tejada, B., Schäffer, L., Papadia, A., Radan, A. P., Todesco-Bernasconi, M., Ville, Y., Voekt, C. A., Eggel-Hort, B., Capoccia-Brugger, R., Johann, S., Grawe, C., Defert, S., Mottet, N., Kahlert, C. R., Garabedian, C., Sentilhes, L., Weber, B., Leu, S., Bassler, D., Lepigeon, K., Winterfeld, U., Panchaud, A., Baud, D. & French and Swiss COVI-PREG group. Maternal and perinatal outcomes following pre-Delta, Delta, and Omicron SARS-CoV-2 variants infection among unvaccinated pregnant women in France and Switzerland: a prospective cohort study using the COVI-PREG registry.Lancet Reg Health Eur 100569 (2023). doi:10.1016/j.lanepe.2022.100569

17. Smith, E. R., Oakley, E., Grandner, G. W., Rukundo, G., Farooq, F., Ferguson, K., Baumann, S., Waldorf, K. A., Afshar, Y., Ahlberg, M., Ahmadzia, H., Akelo, V., Aldrovandi, G., Bevilacqua, E., Bracero, N., Brandt, J. S., Broutet, N., Carrillo, J., Conry, J., Cosmi, E., Crispi, F., Crovetto, F., Gil, M. D. M., Delgado-López, C., Divakar, H., Driscoll, A. J., Favre, G., Buhigas, I. F., Flaherman, V., Gale, C., Godwin, C. L., Gottlieb, S., Gratacós, E., He, S., Hernandez, O., Jones, S., Joshi, S., Kalafat, E., Khagayi, S., Knight, M., Kotloff, K., Lanzone, A., Longo, V. L., Le Doare, K., Lees, C., Litman, E., Lokken, E. M., Madhi, S. A., Magee, L. A., Martinez-Portilla, R. J., Metz, T. D., Miller, E. S., Money, D., Moungmaithong, S., Mullins, E., Nachega, J. B., Nunes, M. C., Onyango, D., Panchaud, A., Poon, L. C., Raiten, D., Regan, L., Sahota, D., Sakowicz, A., Sanin-Blair, J., Stephansson, O., Temmerman, M., Thorson, A., Thwin, S. S., Tippett Barr, B. A., Tolosa, J. E., Tug, N., Valencia-Prado, M., Visentin, S., von Dadelszen, P., Whitehead, C., Wood, M., Yang, H., Zavala, R. & Tielsch, J. M. Clinical risk factors of adverse outcomes among women with COVID-19 in the pregnancy and postpartum period: A sequential, prospective meta-analysis. Am. J. Obstet. Gynecol. (2022). doi:10.1016/j.ajog.2022.08.038

18. Maternal deaths. at <https://www.who.int/data/gho/indicator-metadata-registry/imr-details/4622>

19. Barfield, W. D. & COMMITTEE ON FETUS AND NEWBORN. Standard Terminology for Fetal, Infant, and Perinatal Deaths. Pediatrics 137, (2016).

20. Newborn mortality. at <https://www.who.int/news-room/fact-sheets/detail/levels-and-trends-in-child-mortality-report-2021>

21. Tavares Da Silva, F., Gonik, B., McMillan, M., Keech, C., Dellicour, S., Bhange, S., Tila, M., Harper, D. M., Woods, C., Kawai, A. T., Kochhar, S., Munoz, F. M. & Brighton Collaboration Stillbirth Working Group. Stillbirth: Case definition and guidelines for data collection, analysis, and presentation of maternal immunization safety data. Vaccine 34, 6057–6068 (2016).

22. Pathirana, J., Muñoz, F. M., Abbing-Karahagopian, V., Bhat, N., Harris, T., Kapoor, A., Keene, D. L., Mangili, A., Padula, M. A., Pande, S. L., Pool, V., Pourmalek, F., Varricchio, F., Kochhar, S., Cutland, C. L. & Brighton Collaboration Neonatal Death Working Group. Neonatal death: Case definition & guidelines for data collection, analysis, and presentation of immunization safety data. Vaccine 34, 6027–6037 (2016).

23. Tracking SARS-CoV-2 variants. at <https://www.who.int/activities/tracking-SARS-CoV-2-variants>

24. Grannis, S. J., Rowley, E. A., Ong, T. C., Stenehjem, E., Klein, N. P., DeSilva, M. B., Naleway, A. L., Natarajan, K., Thompson, M. G. & VISION Network. Interim Estimates of COVID-19 Vaccine Effectiveness Against COVID-19-Associated Emergency Department or Urgent Care Clinic Encounters and Hospitalizations Among Adults During SARS-CoV-2 B.1.617.2 (Delta) Variant Predominance -Nine States, June-August 2021. MMWR Morb. Mortal. Wkly. Rep. 70, 1291–1293 (2021).

25. Hadfield, J., Megill, C., Bell, S. M., Huddleston, J., Potter, B., Callender, C., Sagulenko, P., Bedford, T. & Neher, R. A. Nextstrain: real-time tracking of pathogen evolution. Bioinformatics 34, 4121–4123 (2018).

26. Balduzzi, S., Rücker, G. & Schwarzer, G. How to perform a meta-analysis with R: a practical tutorial. Evid. Based. Ment. Health 22, 153–160 (2019).

27. Fleiss, J. L. The statistical basis of meta-analysis. Stat. Methods Med. Res. 2, 121–145 (1993).

28. R Core Team. A language and environment for statistical computing. R Foundation for Statistical Computing. (2022). at <https://www.R-project.org/>

29. Litman, E. A., Yin, Y., Nelson, S. J., Capbarat, E., Kerchner, D. & Ahmadzia, H. K. Adverse perinatal outcomes in a large United States birth cohort during the COVID-19 pandemic. Am J Obstet Gynecol MFM 4, 100577 (2022).

30. Jafari, M., Pormohammad, A., Sheikh Neshin, S. A., Ghorbani, S., Bose, D., Alimohammadi, S., Basirjafari, S., Mohammadi, M., Rasmussen-Ivey, C., Razizadeh, M. H., Nouri-Vaskeh, M. & Zarei, M. Clinical characteristics and outcomes of pregnant women with COVID-19 and comparison with control patients: A systematic review and meta-analysis. Rev. Med. Virol. 31, 1–16 (2021).

31. Rangchaikul, P. & Venketaraman, V. SARS-CoV-2 and the Immune Response in Pregnancy with Delta Variant Considerations. Infect. Dis. Rep. 13, 993–1008 (2021).

32. Shanes, E. D., Miller, E. S., Otero, S., Ebbott, R., Aggarwal, R., Willnow, A. S., Ozer, E. A., Mithal, L. B. & Goldstein, J. A. Placental Pathology After SARS-CoV-2 Infection in the Pre-Variant of Concern, Alpha / Gamma, Delta, or Omicron Eras. Int. J. Surg. Pathol. 10668969221102534 (2022). doi:10.1177/10668969221102534

33. Aleem, A., Akbar Samad, A. B. & Slenker, A. K. in StatPearls (StatPearls Publishing, 2022). at <https://www.ncbi.nlm.nih.gov/pubmed/34033342>

34. Menni, C., Valdes, A. M., Polidori, L., Antonelli, M., Penamakuri, S., Nogal, A., Louca, P., May, A., Figueiredo, J. C., Hu, C., Molteni, E., Canas, L., Österdahl, M. F., Modat, M., Sudre, C. H., Fox, B., Hammers, A., Wolf, J., Capdevila, J., Chan, A. T., David, S. P., Steves, C. J., Ourselin, S. & Spector, T. D. Symptom prevalence, duration, and risk of hospital admission in individuals infected with SARS-CoV-2 during periods of omicron and delta variant dominance: a prospective observational study from the ZOE COVID Study. Lancet 399, 1618–1624 (2022).

35. Adhikari, E. H., MacDonald, L., SoRelle, J. A., Morse, J., Pruszynski, J. & Spong, C. Y. COVID-19 Cases and Disease Severity in Pregnancy and Neonatal Positivity Associated With Delta (B.1.617.2) and Omicron (B.1.1.529) Variant Predominance. JAMA 327, 1500–1502 (2022).

36. Adhikari, E. H., SoRelle, J. A., McIntire, D. D. & Spong, C. Y. Increasing severity of COVID-19 in pregnancy with Delta (B.1.617.2) variant surge. Am. J. Obstet. Gynecol. 226, 149–151 (2022).

37. Mndala, L., Monk, E. J. M., Phiri, D., Riches, J., Makuluni, R., Gadama, L., Kachale, F., Bilesi, R., Mbewe, M., Likaka, A., Chapuma, C., Kumwenda, M., Maseko, B., Ndamala, C., Kuyere, A., Munthali, L., Henrion, M. Y. R., Masesa, C. & Lissauer, D. Comparison of maternal and neonatal outcomes of COVID-19 before and after SARS-CoV-2 omicron emergence in maternity facilities in Malawi (MATSurvey): data from a national maternal surveillance platform. Lancet Glob Health 10, e1623–e1631 (2022).

38. National center for biotechnology information. at <http://www.ncbi.nlm.nih.gov>

39. GISAID - gisaid.org. at <http://www.gisaid.org>

40. Bacterial and viral bioinformatics resource center. at <http://www.viprbrc.org>

41. Medic, S., Anastassopoulou, C., Lozanov-Crvenković, Z., Vuković, V., Dragnić, N., Petrović, V., Ristić, M., Pustahija, T., Gojković, Z., Tsakris, A. & Ioannidis, J. P. A. Risk and severity of SARS-CoV-2 reinfections during 2020-2022 in Vojvodina, Serbia: A population-level observational study. Lancet Reg Health Eur 20, 100453 (2022).

42. Altarawneh, H. N., Chemaitelly, H., Hasan, M. R., Ayoub, H. H., Qassim, S., AlMukdad, S., Coyle, P., Yassine, H. M., Al-Khatib, H. A., Benslimane, F. M., Al-Kanaani, Z., Al-Kuwari, E., Jeremijenko, A., Kaleeckal, A. H., Latif, A. N., Shaik, R. M., Abdul-Rahim, H. F., Nasrallah, G. K., Al-Kuwari, M. G., Butt, A. A., Al-Romaihi, H. E., Al-Thani, M. H., Al-Khal, A., Bertollini, R., Tang, P. & Abu-Raddad, L. J. Protection against the Omicron Variant from Previous SARS-CoV-2 Infection. N. Engl. J. Med. 386, 1288–1290 (2022).

43. da Costa, V. G., Saivish, M. V., Santos, D. E. R., de Lima Silva, R. F. & Moreli, M. L. Comparative epidemiology between the 2009 H1N1 influenza and COVID-19 pandemics. J. Infect. Public Health 13, 1797–1804 (2020).

44. P S. & Dhandapani N S. K. Evaluation of Pregnancy, Younger Age, and Old Age as Independent Risk Factors for Poor Hospitalization Outcomes in Influenza A (H1N1)pdm09 Virus a Decade After the Pandemic. Cureus 12, e11762 (2020).

45. Laake, I., Tunheim, G., Robertson, A. H., Hungnes, O., Waalen, K., Håberg, S. E., Mjaaland, S. & Trogstad, L. Risk of pregnancy complications and adverse birth outcomes after maternal A(H1N1)pdm09 influenza: a Norwegian population-based cohort study. BMC Infect. Dis. 18, 525 (2018).

