## Supplemental Tables S1 and S2 for "Risk of Adverse Maternal and Fetal Outcomes Associated with COVID-19 Variants of Concern: A Sequential Prospective Meta-Analysis"

**^#^PMA collaborators**

**Adverse Pregnancy Outcomes Associated with COVID-19 Infection (ANCOV) Kenya Study (Akelo, Tippett Barr, et al., 2021)**

1. Richard Omore, Kenya Medical Research Institute, Center for Global Health Research [KEMRI-CGHR], Kisumu, Kenya
2. Gregory Ouma, KEMRI-CGHR, Kisumu, Kenya
3. Clayton Onyango, US Centers for Disease Control and Prevention, Kisumu, Kenya
4. Kephas Otieno, KEMRI-CGHR, Kisumu, Kenya
5. Zacchaeus Abaja Were, KEMRI-CGHR, Kisumu, Kenya
6. Joyce Were, KEMRI-CGHR, Kisumu, Kenya

**^#^COVI-PREG International Registry**

1. Emeline Maisonneuve, Institute of Primary Health Care (BIHAM), University of Bern, Bern, Switzerland.
2. Christophe Poncelet, Department of Obstetrics and Gynecology, Pontoise Hospital, Pontoise, France
3. Begoña Martinez de Tejada, Obstetrics Division, Department of Pediatrics, Gynecology, and Obstetrics Geneva University Hospitals & Faculty of Medicine, University of Geneva, Geneva, Switzerland.
4. Thibaud Quibel, Department of Obstetrics and Gynecology, Poissy-Saint Germain Hospital, Poissy, France
5. Cécile Monod, Department of Obstetrics, Basel University Hospital, Basel, Switzerland

COVI-Preg International Registry: please add up to 5 co-authors from France and Switzerland

**Hong Kong, SAR, China (Poon et al., 2021)**

1. Florrie NY Yu, Department of Obstetrics and Gynaecology, Queen Elizabeth Hospital, Hong Kong SAR, China
2. Choi Wah Kong, Department of Obstetrics and Gynaecology, United Christian Hospital, Hong Kong SAR, China
3. Tsz Kin Lo, Department of Obstetrics and Gynaecology, Princess Margaret Hospital, Hong Kong SAR, China
4. Po Lam So, Department of Obstetrics and Gynaecology, Tuen Mun Hospital, Hong Kong SAR, China
5. Wing Cheong Leung, Department of Obstetrics and Gynaecology, Kwong Wah Hospital, Hong Kong SAR, China

**Rome Hospital Study (Bevilacqua, Laurita Longo, et al., 2020)**

1. Federica Meli, Department of Women's and Child Health Sciences and Public Health, IRCCS A. Gemelli University Polyclinic Foundation, Rome, Italy
2. Giulia Bonanni, Department of Women's and Child Health Sciences and Public Health, IRCCS A. Gemelli University Polyclinic Foundation, Rome, Italy
3. Federica Romanzi, Department of Women's and Child Health Sciences and Public Health, IRCCS A. Gemelli University Polyclinic Foundation, Rome, Italy
4. Eleonora Torcia, Department of Women's and Child Health Sciences and Public Health, IRCCS A. Gemelli University Polyclinic Foundation, Rome, Italy
5. Chiara di Ilio, Department of Women's and Child Health Sciences and Public Health, IRCCS A. Gemelli University Polyclinic Foundation, Rome, Italy

**Madrid Hospital Registry Study (Gil, Fernandez Buhigas, 2021)**

1. Adriana Aquise, Department of Obstetrics and Gynecology, Hospital Universitario de Torrejón, Torrejón de Ardoz, Madrid, Spain
2. María Nieves Rayo, Department of Obstetrics and Gynecology, Hospital Universitario de Torrejón, Torrejón de Ardoz, Madrid, Spain
3. Belén Santacruz, Department of Obstetrics and Gynecology, Hospital Universitario de Torrejón, Torrejón de Ardoz, Madrid, Spain
4. Lorena Gonzalez-Gea, Department of Obstetrics and Gynecology, Hospital Universitario de Torrejón, Torrejón de Ardoz, Madrid, Spain
5. Sheila Laiseca, Department of Obstetrics and Gynecology, Hospital Universitario de Torrejón, Torrejón de Ardoz, Madrid, Spain

**The Puerto Rico COVID-19 Antenatal Sentinel Surveillance System (CASSS) (Bracero, Valencia, Delgado-Lopez, 2021)**

1. Leishla Nieves Ferrer, Surveillance for Emerging Threats to Mothers and Babies, Puerto Rico Department of Health, San Juan, Puerto Rico
2. Mariam Marcano Huertas, Surveillance for Emerging Threats to Mothers and Babies, Puerto Rico Department of Health, San Juan, Puerto Rico
3. Glorimar Meléndez Rosario, Surveillance for Emerging Threats to Mothers and Babies, Puerto Rico Department of Health, San Juan, Puerto Rico
4. Nadjarie Aviles Ramos, Surveillance for Emerging Threats to Mothers and Babies, Puerto Rico Department of Health, San Juan, Puerto Rico
5. Stephanie Vicente González, Surveillance for Emerging Threats to Mothers and Babies, Puerto Rico Department of Health, San Juan, Puerto Rico

***Corresponding Author**

Fouzia Farooq

The George Washington University

Milken Institute School of Public of Public Health

950 New Hampshire Ave, NW, Suite 400

Washington, DC 20052 USA

**Conflicts of Interest:**

Authors have no conflicts of interest to disclose.

**Supplementary Tables**

**Table S1. Random effect relative risk (RR) estimates with 95% confidence intervals (CI) and number of studies included in each analysis (dominant strain threshold ≥60%)**

A. **COVID-19 Severity Outcomes**

| **Outcome** | **Reference Strain** | **Pre-alpha** | **Alpha** | **Beta** | **Delta** | **Epsilon** | **Eta** | **Omicron** | **Mixed** |
| --- | --- | --- | --- | --- | --- | --- | --- | --- | --- |
| **Hospitalization*** | **Pre-alpha** | Reference | 1.41 (0.76, 2.63), n=4 | 0.92 (0.54, 1.54), n=1 | 1.12 (0.48, 2.58), n=2 | 1.57 (1.15, 2.14), n=1 | 1.21 (0.17, 8.66), n=1 | 0.99 (0.87, 1.11), n=2 | 1.13 (0.95, 1.33), n=5 |
|  | **Alpha** | 0.71 (0.38, 1.32), n=4 | Reference | 1.00 (0.49, 2.05), n=1 | 0.56 (0.23, 1.35), n=2 | n=0 | 0.36 (0.05, 2.68), n=1 | 0.41 (0.08, 1.98), n=2 | 0.70 (0.39, 1.25), n=4 |
|  | **Beta** | 1.09 (0.65, 1.84), n=1 | 1.00 (0.49, 2.05), n=1 | Reference | n=0 | n=0 | n=0 | 1.10 (0.66, 1.83), n=1 | 0.90 (0.35, 2.32), n=1 |
|  | **Delta** | 0.89 (0.39, 2.07), n=2 | 1.80 (0.74, 4.37), n=2 | n=0 | Reference | n=0 | 0.62 (0.08, 4.77), n=1 | 0.18 (0.05, 0.73), n=1 | 0.72 (0.28, 1.89), n=2 |
|  | **Epsilon** | 0.64 (0.47, 0.87), n=1 | n=0 | n=0 | n=0 | Reference | n=0 | n=0 | 0.73 (0.51, 1.03), n=1 |
|  | **Eta** | 0.83 (0.12, 5.93), n=1 | 2.77 (0.37, 20.53), n=1 | n=0 | 1.62 (0.21, 12.55), n=1 | n=0 | Reference | 0.30 (0.03, 2.62), n=1 | 1.00 (0.13, 7.43), n=1 |
|  | **Omicron** | 1.01 (0.90, 1.15), n=2 | 2.47 (0.51, 12.02), n=2 | 0.91 (0.55, 1.51), n=1 | 5.50 (1.38, 21.92), n=1 | n=0 | 3.39 (0.38, 30.10), n=1 | Reference | 1.33 (0.55, 3.22), n=2 |
|  | **Mixed** | 0.89 (0.75, 1.05), n=5 | 1.43 (0.80, 2.56), n=4 | 1.11 (0.43, 2.86), n=1 | 1.38 (0.53, 3.61), n=2 | 1.38 (0.97, 1.96), n=1 | 1.00 (0.13, 7.43), n=1 | 0.75 (0.31, 1.82), n=2 | Reference |
| **ICU admission** | **Pre-alpha** | Reference | 1.39 (0.77, 2.51), n=4 | 3.11 (0.16, 60.70), n=1 | 2.14 (0.97, 4.74), n=3 | 0.47 (0.07, 3.28), n=1 | 12.08 (0.81, 181.34), n=1 | 0.67 (0.22, 2.04), n=4 | 1.27 (0.77, 2.07), n=5 |
|  | **Alpha** | 0.72 (0.40, 1.29), n=4 | Reference | n=0 | 1.28 (0.60, 2.72), n=3 | n=0 | 6.23 (0.27, 142.73), n=1 | 0.38 (0.14, 1.06), n=4 | 0.89 (0.26, 3.09), n=3 |
|  | **Beta** | 0.32 (0.02, 6.27), n=1 | n=0 | Reference | n=0 | n=0 | n=0 | 0.10 (0.01, 1.98), n=1 | n=0 |
|  | **Delta** | 0.47 (0.21, 1.03), n=3 | 0.78 (0.37, 1.66), n=3 | n=0 | Reference | n=0 | 3.08 (0.21, 45.62), n=1 | 0.34 (0.15, 0.73), n=3 | 0.53 (0.18, 1.55), n=3 |
|  | **Epsilon** | 2.15 (0.30, 15.14), n=1 | n=0 | n=0 | n=0 | Reference | n=0 | n=0 | 2.35 (0.31, 17.83), n=1 |
|  | **Eta** | 0.08 (0.01, 1.24), n=1 | 0.16 (0.01, 3.68), n=1 | n=0 | 0.32 (0.02, 4.80), n=1 | n=0 | Reference | 0.10 (0.01, 1.47), n=1 | 0.43 (0.05, 3.79), n=1 |
|  | **Omicron** | 1.48 (0.49, 4.49), n=4 | 2.63 (0.95, 7.30), n=4 | 10.00 (0.50, 198.21), n=1 | 2.98 (1.37, 6.47), n=3 | n=0 | 10.17 (0.68, 152.44), n=1 | Reference | 1.82 (0.68, 4.86), n=4 |
|  | **Mixed** | 0.79 (0.48, 1.29), n=5 | 1.12 (0.32, 3.89), n=3 | n=0 | 1.88 (0.64, 5.51), n=3 | 0.43 (0.06, 3.23), n=1 | 2.33 (0.26, 20.66), n=1 | 0.55 (0.21, 1.47), n=4 | Reference |
| **Critical care** | **Pre-alpha** | Reference | 1.32 (0.73, 2.37), n=4 | n=0 | 1.91 (0.98, 3.73), n=4 | 0.39 (0.06, 2.74), n=1 | 6.04 (0.59, 61.92), n=1 | 0.71 (0.20, 2.46), n=3 | 1.06 (0.65, 1.73), n=5 |
|  | **Alpha** | 0.76 (0.42, 1.37), n=4 | Reference | n=0 | 1.39 (0.67, 2.87), n=3 | n=0 | 6.23 (0.27, 142.73), n=1 | 0.47 (0.15, 1.48), n=3 | 0.89 (0.26, 3.09), n=3 |
|  | **Beta** | n=0 | n=0 | Reference | n=0 | n=0 | n=0 | n=0 | n=0 |
|  | **Delta** | 0.52 (0.27, 1.02), n=4 | 0.72 (0.35, 1.49), n=3 | n=0 | Reference | n=0 | 3.08 (0.21, 45.62), n=1 | 0.34 (0.16, 0.73), n=3 | 0.52 (0.18, 1.52), n=3 |
|  | **Epsilon** | 2.57 (0.36, 18.10), n=1 | n=0 | n=0 | n=0 | Reference | n=0 | n=0 | 2.35 (0.31, 17.83), n=1 |
|  | **Eta** | 0.17 (0.02, 1.70), n=1 | 0.16 (0.01, 3.68), n=1 | n=0 | 0.32 (0.02, 4.80), n=1 | n=0 | Reference | 0.10 (0.01, 1.47), n=1 | 0.43 (0.05, 3.79), n=1 |
|  | **Omicron** | 1.41 (0.41, 4.88), n=3 | 2.12 (0.67, 6.66), n=3 | n=0 | 2.94 (1.38, 6.29), n=3 | n=0 | 10.17 (0.68, 152.44), n=1 | Reference | 1.28 (0.45, 3.60), n=3 |
|  | **Mixed** | 0.94 (0.58, 1.53), n=5 | 1.12 (0.32, 3.89), n=3 | n=0 | 1.92 (0.66, 5.62), n=3 | 0.43 (0.06, 3.23), n=1 | 2.33 (0.26, 20.66), n=1 | 0.78 (0.28, 2.20), n=3 | Reference |
| **Ventilation** | **Pre-alpha** | Reference | 2.10 (0.93, 4.76), n=5 | 3.11 (0.16, 60.70), n=1 | 2.70 (1.14, 6.41), n=4 | 0.38 (0.02, 6.02), n=1 | 6.04 (0.59, 61.92), n=1 | 0.70 (0.26, 1.85), n=4 | 1.25 (0.39, 4.03), n=6 |
|  | **Alpha** | 0.48 (0.21, 1.07), n=5 | Reference | n=0 | 1.62 (0.65, 4.02), n=2 | n=0 | 6.23 (0.27, 142.73), n=1 | 0.30 (0.11, 0.81), n=4 | 0.81 (0.19, 3.40), n=3 |
|  | **Beta** | 0.32 (0.02, 6.27), n=1 | n=0 | Reference | n=0 | n=0 | n=0 | 0.10 (0.01, 1.98), n=1 | n=0 |
|  | **Delta** | 0.37 (0.16, 0.88), n=4 | 0.62 (0.25, 1.53), n=2 | n=0 | Reference | n=0 | 8.77 (0.38, 202.21), n=1 | 0.23 (0.08, 0.65), n=3 | 0.58 (0.15, 2.25), n=3 |
|  | **Epsilon** | 2.65 (0.17, 42.22), n=1 | n=0 | n=0 | n=0 | Reference | n=0 | n=0 | 0.99 (0.05, 20.39), n=1 |
|  | **Eta** | 0.17 (0.02, 1.70), n=1 | 0.16 (0.01, 3.68), n=1 | n=0 | 0.11 (0.00, 2.63), n=1 | n=0 | Reference | 0.10 (0.01, 1.47), n=1 | 0.14 (0.01, 2.14), n=1 |
|  | **Omicron** | 1.43 (0.54, 3.78), n=4 | 3.31 (1.24, 8.84), n=4 | 10.00 (0.50, 198.21), n=1 | 4.31 (1.55, 12.01), n=3 | n=0 | 10.17 (0.68, 152.44), n=1 | Reference | 2.99 (0.85, 10.51), n=4 |
|  | **Mixed** | 0.80 (0.25, 2.57), n=6 | 1.23 (0.29, 5.13), n=3 | n=0 | 1.72 (0.44, 6.69), n=3 | 1.01 (0.05, 20.93), n=1 | 7.00 (0.47, 104.68), n=1 | 0.33 (0.10, 1.18), n=4 | Reference |
| **Pneumonia** | **Pre-alpha** | Reference | 1.13 (0.55, 2.31), n=4 | n=0 | 0.73 (0.29, 1.80), n=4 | 1.61 (0.94, 2.73), n=1 | 0.76 (0.11, 5.22), n=1 | 0.30 (0.10, 0.89), n=3 | 1.32 (1.04, 1.69), n=5 |
|  | **Alpha** | 0.89 (0.43, 1.82), n=4 | Reference | n=0 | 0.65 (0.23, 1.84), n=4 | n=0 | 0.54 (0.07, 4.34), n=1 | 0.32 (0.05, 1.86), n=3 | 0.91 (0.37, 2.21), n=3 |
|  | **Beta** | n=0 | n=0 | Reference | n=0 | n=0 | n=0 | n=0 | n=0 |
|  | **Delta** | 1.37 (0.56, 3.39), n=4 | 1.53 (0.54, 4.32), n=4 | n=0 | Reference | n=0 | 0.77 (0.10, 6.25), n=1 | 0.20 (0.05, 0.71), n=3 | 1.41 (0.62, 3.19), n=4 |
|  | **Epsilon** | 0.62 (0.37, 1.06), n=1 | n=0 | n=0 | n=0 | Reference | n=0 | n=0 | 0.81 (0.45, 1.46), n=1 |
|  | **Eta** | 1.32 (0.19, 9.15), n=1 | 1.85 (0.23, 14.80), n=1 | n=0 | 1.30 (0.16, 10.51), n=1 | n=0 | Reference | 0.10 (0.01, 1.47), n=1 | 1.29 (0.18, 9.27), n=1 |
|  | **Omicron** | 3.33 (1.13, 9.84), n=3 | 3.14 (0.54, 18.36), n=3 | n=0 | 5.07 (1.41, 18.25), n=3 | n=0 | 10.17 (0.68, 152.44), n=1 | Reference | 7.81 (2.40, 25.43), n=3 |
|  | **Mixed** | 0.75 (0.59, 0.97), n=5 | 1.10 (0.45, 2.69), n=3 | n=0 | 0.71 (0.31, 1.61), n=4 | 1.23 (0.68, 2.21), n=1 | 0.78 (0.11, 5.61), n=1 | 0.13 (0.04, 0.42), n=3 | Reference |

*Hospitalization was categorized as follows:

Martinez-Portilla, 2021: Clearly defined hospitalization for COVID-19 | Gill, Fernandez Buhigas, 2021: Clearly defined hospitalization for COVID-19 | Bevilacqua, Laurita Longo, 2020: Hospitalization for COVID-19 included those for L&D as well as pregnancy-related preterm labor/births. | Akelo, Tippett Barr 2021: Clearly defined hospitalization for COVID-19 | Poon 2021: Hospitalization for COVD-19 included any hospital admission

B. **Maternal Morbidity & Mortality**

| **Outcome** | **Reference Strain** | **Pre-alpha** | **Alpha** | **Beta** | **Delta** | | | **Epsilon** | **Eta** | **Omicron** | **Mixed** |
| --- | --- | --- | --- | --- | --- | --- | --- | --- | --- | --- | --- |
| **Pregnancy-related death** | **Pre-alpha** | Reference | 0.99 (0.09, 10.82), n=1 | n=0 | | 0.73 (0.04, 15.04), n=1 | 0.86 (0.12, 6.06), n=1 | | n=0 | 0.28 (0.01, 5.86), n=1 | 0.40 (0.07, 2.15), n=2 |
|  | **Alpha** | 1.01 (0.09, 11.08), n=1 | Reference | n=0 | | 0.62 (0.03, 14.97), n=1 | n=0 | | n=0 | 0.24 (0.01, 5.83), n=1 | 2.46 (0.10, 58.71), n=1 |
|  | **Beta** | n=0 | n=0 | Reference | | n=0 | n=0 | | n=0 | n=0 | n=0 |
|  | **Delta** | 1.37 (0.07, 28.35), n=1 | 1.62 (0.07, 39.45), n=1 | n=0 | | Reference | n=0 | | n=0 | n=0 | n=0 |
|  | **Epsilon** | 1.17 (0.17, 8.26), n=1 | n=0 | n=0 | | n=0 | Reference | | n=0 | n=0 | 0.20 (0.01, 3.09), n=1 |
|  | **Eta** | n=0 | n=0 | n=0 | | n=0 | n=0 | | Reference | n=0 | n=0 |
|  | **Omicron** | 3.54 (0.17, 73.34), n=1 | 4.18 (0.17, 102.04), n=1 | n=0 | | n=0 | n=0 | | n=0 | Reference | n=0 |
|  | **Mixed** | 2.50 (0.47, 13.42), n=2 | 0.41 (0.02, 9.67), n=1 | n=0 | | n=0 | 5.12 (0.32, 81.14), n=1 | | n=0 | n=0 | Reference |
| **Placental abruption** | **Pre-alpha** | Reference | 2.77 (0.54, 14.16), n=3 | 3.11 (0.16, 60.70), n=1 | | 0.48 (0.05, 4.63), n=2 | n=0 | | n=0 | 0.37 (0.05, 2.92), n=2 | 4.68 (0.87, 25.14), n=3 |
|  | **Alpha** | 0.36 (0.07, 1.84), n=3 | Reference | n=0 | | 0.18 (0.02, 1.76), n=2 | n=0 | | n=0 | 0.15 (0.02, 1.33), n=2 | 1.71 (0.22, 13.56), n=2 |
|  | **Beta** | 0.32 (0.02, 6.27), n=1 | n=0 | Reference | | n=0 | n=0 | | n=0 | 0.10 (0.01, 1.98), n=1 | n=0 |
|  | **Delta** | 2.07 (0.22, 19.77), n=2 | 5.41 (0.57, 51.51), n=2 | n=0 | | Reference | n=0 | | n=0 | n=0 | 23.21 (0.96, 558.37), n=1 |
|  | **Omicron** | 2.74 (0.34, 21.89), n=2 | 6.66 (0.75, 59.01), n=2 | 10.00 (0.50, 198.21), n=1 | | n=0 | n=0 | | n=0 | Reference | 15.00 (0.84, 266.57), n=1 |
|  | **Mixed** | 0.21 (0.04, 1.15), n=3 | 0.58 (0.07, 4.64), n=2 | n=0 | | 0.04 (0.00, 1.04), n=1 | n=0 | | n=0 | 0.07 (0.00, 1.18), n=1 | Reference |
| **Preterm labor** | **Pre-alpha** | Reference | 0.63 (0.27, 1.51), n=4 | 1.04 (0.07, 14.95), n=1 | | 1.56 (0.77, 3.14), n=3 | n=0 | | 2.25 (0.11, 44.35), n=1 | 0.80 (0.42, 1.52), n=3 | 2.40 (1.00, 5.76), n=4 |
|  | **Alpha** | 1.58 (0.66, 3.77), n=4 | Reference | n=0 | | 2.18 (0.75, 6.30), n=3 | n=0 | | n=0 | 1.19 (0.43, 3.33), n=3 | 2.80 (0.72, 10.85), n=3 |
|  | **Beta** | 0.96 (0.07, 13.90), n=1 | n=0 | Reference | | n=0 | n=0 | | n=0 | 0.63 (0.05, 8.53), n=1 | n=0 |
|  | **Delta** | 0.64 (0.32, 1.30), n=3 | 0.46 (0.16, 1.33), n=3 | n=0 | | Reference | n=0 | | 0.97 (0.04, 22.47), n=1 | 0.78 (0.27, 2.22), n=2 | 1.61 (0.70, 3.70), n=3 |
|  | **Eta** | 0.45 (0.02, 8.79), n=1 | n=0 | n=0 | | 1.03 (0.04, 23.67), n=1 | n=0 | | Reference | 0.95 (0.05, 16.70), n=1 | 0.46 (0.02, 10.67), n=1 |
|  | **Omicron** | 1.25 (0.66, 2.35), n=3 | 0.84 (0.30, 2.34), n=3 | 1.58 (0.12, 21.28), n=1 | | 1.29 (0.45, 3.69), n=2 | n=0 | | 1.05 (0.06, 18.45), n=1 | Reference | 1.05 (0.29, 3.75), n=3 |
|  | **Mixed** | 0.42 (0.17, 1.00), n=4 | 0.36 (0.09, 1.39), n=3 | n=0 | | 0.62 (0.27, 1.43), n=3 | n=0 | | 2.18 (0.09, 50.71), n=1 | 0.96 (0.27, 3.43), n=3 | Reference |
| **Preterm labor (COVID-19 onset <37w)** | **Pre-alpha** | Reference | 0.88 (0.58, 1.34), n=2 | n=0 | | 1.40 (0.76, 2.61), n=2 | n=0 | | n=0 | 0.99 (0.54, 1.81), n=2 | 0.96 (0.53, 1.75), n=2 |
|  | **Alpha** | 1.14 (0.75, 1.73), n=2 | Reference | n=0 | | 1.44 (0.83, 2.50), n=2 | n=0 | | n=0 | 1.03 (0.64, 1.65), n=2 | 0.92 (0.44, 1.92), n=2 |
|  | **Delta** | 0.71 (0.38, 1.32), n=2 | 0.69 (0.40, 1.20), n=2 | n=0 | | Reference | n=0 | | n=0 | 0.73 (0.46, 1.14), n=2 | 0.56 (0.28, 1.10), n=2 |
|  | **Omicron** | 1.01 (0.55, 1.84), n=2 | 0.97 (0.61, 1.56), n=2 | n=0 | | 1.38 (0.88, 2.17), n=2 | n=0 | | n=0 | Reference | 0.77 (0.41, 1.43), n=2 |
|  | **Mixed** | 1.04 (0.57, 1.90), n=2 | 1.09 (0.52, 2.27), n=2 | n=0 | | 1.79 (0.91, 3.51), n=2 | n=0 | | n=0 | 1.30 (0.70, 2.42), n=2 | Reference |
| **Hemorrhage** | **Pre-alpha** | Reference | 2.00 (1.05, 3.83), n=3 | 6.75 (0.99, 46.19), n=1 | | 1.19 (0.57, 2.46), n=2 | n=0 | | 2.10 (0.87, 5.08), n=1 | 1.17 (0.71, 1.94), n=2 | 1.68 (1.01, 2.78), n=3 |
|  | **Alpha** | 0.50 (0.26, 0.96), n=3 | Reference | 3.00 (0.19, 47.96), n=1 | | 0.57 (0.25, 1.30), n=2 | n=0 | | 1.08 (0.40, 2.90), n=1 | 0.61 (0.31, 1.17), n=2 | 0.83 (0.43, 1.61), n=2 |
|  | **Beta** | 0.15 (0.02, 1.01), n=1 | 0.33 (0.02, 5.33), n=1 | Reference | | n=0 | n=0 | | n=0 | 0.02 (0.00, 0.25), n=1 | 0.50 (0.04, 7.10), n=1 |
|  | **Delta** | 0.84 (0.41, 1.74), n=2 | 1.75 (0.77, 4.01), n=2 | n=0 | | Reference | n=0 | | 1.76 (0.62, 4.99), n=1 | 1.08 (0.51, 2.30), n=1 | 1.39 (0.67, 2.91), n=2 |
|  | **Eta** | 0.48 (0.20, 1.15), n=1 | 0.92 (0.34, 2.47), n=1 | n=0 | | 0.57 (0.20, 1.61), n=1 | n=0 | | Reference | 0.61 (0.26, 1.47), n=1 | 0.79 (0.33, 1.89), n=1 |
|  | **Omicron** | 0.85 (0.52, 1.41), n=2 | 1.65 (0.85, 3.18), n=2 | 44.50 (4.07, 486.35), n=1 | | 0.92 (0.43, 1.96), n=1 | n=0 | | 1.63 (0.68, 3.90), n=1 | Reference | 1.37 (0.84, 2.25), n=2 |
|  | **Mixed** | 0.60 (0.36, 0.99), n=3 | 1.21 (0.62, 2.35), n=2 | 2.00 (0.14, 28.42), n=1 | | 0.72 (0.34, 1.50), n=2 | n=0 | | 1.27 (0.53, 3.06), n=1 | 0.73 (0.44, 1.19), n=2 | Reference |
| **Embolic disease** | **Pre-alpha** | Reference | 1.80 (0.08, 43.09), n=1 | n=0 | | 1.28 (0.05, 30.82), n=1 | n=0 | | 3.74 (0.16, 87.35), n=1 | 0.40 (0.02, 9.63), n=1 | 0.57 (0.02, 13.90), n=1 |
|  | **Alpha** | 0.55 (0.02, 13.26), n=1 | Reference | n=0 | | n=0 | n=0 | | n=0 | n=0 | n=0 |
|  | **Beta** | n=0 | n=0 | Reference | | n=0 | n=0 | | n=0 | n=0 | n=0 |
|  | **Delta** | 0.78 (0.03, 18.79), n=1 | n=0 | n=0 | | Reference | n=0 | | n=0 | n=0 | n=0 |
|  | **Eta** | 0.27 (0.01, 6.23), n=1 | n=0 | n=0 | | n=0 | n=0 | | Reference | n=0 | n=0 |
|  | **Omicron** | 2.53 (0.10, 61.49), n=1 | n=0 | n=0 | | n=0 | n=0 | | n=0 | Reference | n=0 |
|  | **Mixed** | 1.75 (0.07, 42.40), n=1 | n=0 | n=0 | | n=0 | n=0 | | n=0 | n=0 | Reference |
| **Preeclampsia** | **Pre-alpha** | Reference | 1.45 (0.61, 3.42), n=3 | 1.87 (0.11, 30.49), n=1 | | 1.15 (0.34, 3.94), n=2 | n=0 | | n=0 | 0.95 (0.44, 2.08), n=2 | 2.95 (0.78, 11.12), n=3 |
|  | **Alpha** | 0.69 (0.29, 1.63), n=3 | Reference | n=0 | | 0.56 (0.18, 1.75), n=2 | n=0 | | n=0 | 0.85 (0.32, 2.23), n=2 | 1.26 (0.31, 5.10), n=2 |
|  | **Beta** | 0.54 (0.03, 8.75), n=1 | n=0 | Reference | | n=0 | n=0 | | n=0 | 0.37 (0.03, 5.22), n=1 | n=0 |
|  | **Delta** | 0.87 (0.25, 2.97), n=2 | 1.79 (0.57, 5.63), n=2 | n=0 | | Reference | n=0 | | n=0 | 1.49 (0.32, 6.90), n=1 | 2.38 (0.61, 9.22), n=2 |
|  | **Omicron** | 1.05 (0.48, 2.28), n=2 | 1.18 (0.45, 3.11), n=2 | 2.73 (0.19, 38.86), n=1 | | 0.67 (0.15, 3.09), n=1 | n=0 | | n=0 | Reference | 2.06 (0.42, 10.05), n=2 |
|  | **Mixed** | 0.34 (0.09, 1.28), n=3 | 0.80 (0.20, 3.23), n=2 | n=0 | | 0.42 (0.11, 1.63), n=2 | n=0 | | n=0 | 0.49 (0.10, 2.37), n=2 | Reference |
| **Eclampsia** | **Pre-alpha** | Reference | n=0 | n=0 | | n=0 | n=0 | | n=0 | n=0 | n=0 |
|  | **Alpha** | n=0 | Reference | n=0 | | n=0 | n=0 | | n=0 | n=0 | n=0 |
|  | **Beta** | n=0 | n=0 | Reference | | n=0 | n=0 | | n=0 | n=0 | n=0 |
|  | **Omicron** | n=0 | n=0 | n=0 | | n=0 | n=0 | | n=0 | Reference | n=0 |
|  | **Mixed** | n=0 | n=0 | n=0 | | n=0 | n=0 | | n=0 | n=0 | Reference |
| **HDP (diagnosed at any time)** | **Pre-alpha** | Reference | 1.45 (0.19, 11.23), n=2 | 1.87 (0.11, 30.49), n=1 | | 0.77 (0.04, 15.67), n=1 | n=0 | | 2.25 (0.11, 44.35), n=1 | 0.91 (0.27, 3.10), n=2 | 1.11 (0.15, 8.27), n=2 |
|  | **Alpha** | 0.69 (0.09, 5.34), n=2 | Reference | n=0 | | n=0 | n=0 | | n=0 | 0.59 (0.08, 4.34), n=2 | n=0 |
|  | **Beta** | 0.54 (0.03, 8.75), n=1 | n=0 | Reference | | n=0 | n=0 | | n=0 | 0.37 (0.03, 5.22), n=1 | n=0 |
|  | **Delta** | 1.30 (0.06, 26.54), n=1 | n=0 | n=0 | | Reference | n=0 | | n=0 | 1.54 (0.08, 31.48), n=1 | n=0 |
|  | **Eta** | 0.45 (0.02, 8.79), n=1 | n=0 | n=0 | | n=0 | n=0 | | Reference | 0.53 (0.03, 10.43), n=1 | n=0 |
|  | **Omicron** | 1.10 (0.32, 3.76), n=2 | 1.69 (0.23, 12.35), n=2 | 2.73 (0.19, 38.86), n=1 | | 0.65 (0.03, 13.19), n=1 | n=0 | | 1.89 (0.10, 37.33), n=1 | Reference | 1.37 (0.20, 9.57), n=2 |
|  | **Mixed** | 0.90 (0.12, 6.66), n=2 | n=0 | n=0 | | n=0 | n=0 | | n=0 | 0.73 (0.10, 5.08), n=2 | Reference |
| **HDP (diagnosed at or after COVID-19)** | **Pre-alpha** | Reference | 1.17 (0.40, 3.40), n=3 | 1.04 (0.07, 14.95), n=1 | | 0.69 (0.27, 1.72), n=2 | n=0 | | 2.25 (0.11, 44.35), n=1 | 0.72 (0.27, 1.89), n=2 | 1.66 (0.66, 4.18), n=4 |
|  | **Alpha** | 0.85 (0.29, 2.48), n=3 | Reference | n=0 | | 0.56 (0.15, 2.06), n=1 | n=0 | | n=0 | 0.75 (0.10, 5.40), n=2 | 1.84 (0.50, 6.76), n=2 |
|  | **Beta** | 0.96 (0.07, 13.90), n=1 | n=0 | Reference | | n=0 | n=0 | | n=0 | 0.57 (0.04, 7.71), n=1 | n=0 |
|  | **Delta** | 1.46 (0.58, 3.65), n=2 | 1.80 (0.49, 6.66), n=1 | n=0 | | Reference | n=0 | | n=0 | 1.54 (0.08, 31.48), n=1 | 2.98 (0.92, 9.60), n=1 |
|  | **Eta** | 0.45 (0.02, 8.79), n=1 | n=0 | n=0 | | n=0 | n=0 | | Reference | 0.53 (0.03, 10.43), n=1 | n=0 |
|  | **Omicron** | 1.39 (0.53, 3.68), n=2 | 1.33 (0.19, 9.54), n=2 | 1.76 (0.13, 24.00), n=1 | | 0.65 (0.03, 13.19), n=1 | n=0 | | 1.89 (0.10, 37.33), n=1 | Reference | 1.08 (0.16, 7.40), n=2 |
|  | **Mixed** | 0.60 (0.24, 1.52), n=4 | 0.54 (0.15, 1.99), n=2 | n=0 | | 0.34 (0.10, 1.08), n=1 | n=0 | | n=0 | 0.92 (0.14, 6.28), n=2 | Reference |
| **C-section** | **Pre-alpha** | Reference | 1.33 (1.06, 1.66), n=5 | 2.22 (1.11, 4.46), n=1 | | 1.01 (0.82, 1.25), n=4 | n=0 | | 1.12 (0.48, 2.59), n=1 | 0.89 (0.76, 1.05), n=4 | 1.20 (0.98, 1.48), n=5 |
|  | **Alpha** | 0.75 (0.60, 0.94), n=5 | Reference | 1.00 (0.49, 2.05), n=1 | | 0.83 (0.61, 1.14), n=4 | n=0 | | 0.79 (0.31, 1.97), n=1 | 0.63 (0.50, 0.78), n=4 | 0.98 (0.73, 1.31), n=5 |
|  | **Beta** | 0.45 (0.22, 0.90), n=1 | 1.00 (0.49, 2.05), n=1 | Reference | | n=0 | n=0 | | n=0 | 0.42 (0.24, 0.75), n=1 | 0.90 (0.35, 2.32), n=1 |
|  | **Delta** | 0.99 (0.80, 1.23), n=4 | 1.20 (0.88, 1.64), n=4 | n=0 | | Reference | n=0 | | 0.75 (0.31, 1.81), n=1 | 0.86 (0.67, 1.10), n=3 | 1.00 (0.69, 1.44), n=4 |
|  | **Eta** | 0.89 (0.39, 2.07), n=1 | 1.27 (0.51, 3.18), n=1 | n=0 | | 1.33 (0.55, 3.21), n=1 | n=0 | | Reference | 0.89 (0.38, 2.06), n=1 | 0.96 (0.41, 2.28), n=1 |
|  | **Omicron** | 1.12 (0.95, 1.32), n=4 | 1.60 (1.28, 2.00), n=4 | 2.38 (1.33, 4.25), n=1 | | 1.17 (0.91, 1.50), n=3 | n=0 | | 1.13 (0.48, 2.63), n=1 | Reference | 1.36 (1.08, 1.71), n=4 |
|  | **Mixed** | 0.83 (0.68, 1.02), n=5 | 1.02 (0.77, 1.37), n=5 | 1.11 (0.43, 2.86), n=1 | | 1.00 (0.69, 1.45), n=4 | n=0 | | 1.04 (0.44, 2.45), n=1 | 0.74 (0.58, 0.93), n=4 | Reference |
| **Intrapartum C-section** | **Pre-alpha** | Reference | 1.48 (1.02, 2.15), n=5 | 1.93 (0.42, 8.87), n=1 | | 1.11 (0.77, 1.62), n=4 | n=0 | | 0.65 (0.10, 4.48), n=1 | 0.94 (0.75, 1.19), n=4 | 1.29 (0.94, 1.77), n=5 |
|  | **Alpha** | 0.68 (0.47, 0.98), n=5 | Reference | 0.60 (0.17, 2.07), n=1 | | 0.80 (0.53, 1.20), n=4 | n=0 | | 0.43 (0.06, 3.32), n=1 | 0.58 (0.39, 0.86), n=4 | 0.99 (0.66, 1.50), n=5 |
|  | **Beta** | 0.52 (0.11, 2.38), n=1 | 1.67 (0.48, 5.76), n=1 | Reference | | n=0 | n=0 | | n=0 | 0.54 (0.13, 2.25), n=1 | 1.50 (0.38, 6.00), n=1 |
|  | **Delta** | 0.90 (0.62, 1.30), n=4 | 1.25 (0.83, 1.88), n=4 | n=0 | | Reference | n=0 | | 0.30 (0.04, 2.11), n=1 | 0.85 (0.56, 1.28), n=3 | 1.02 (0.65, 1.60), n=4 |
|  | **Eta** | 1.53 (0.22, 10.51), n=1 | 2.31 (0.30, 17.66), n=1 | n=0 | | 3.33 (0.47, 23.41), n=1 | n=0 | | Reference | 1.67 (0.24, 11.49), n=1 | 2.00 (0.29, 13.87), n=1 |
|  | **Omicron** | 1.06 (0.84, 1.34), n=4 | 1.73 (1.16, 2.58), n=4 | 1.85 (0.44, 7.73), n=1 | | 1.18 (0.78, 1.78), n=3 | n=0 | | 0.60 (0.09, 4.11), n=1 | Reference | 1.36 (0.97, 1.92), n=4 |
|  | **Mixed** | 0.78 (0.57, 1.07), n=5 | 1.01 (0.67, 1.52), n=5 | 0.67 (0.17, 2.67), n=1 | | 0.98 (0.63, 1.53), n=4 | n=0 | | 0.50 (0.07, 3.47), n=1 | 0.73 (0.52, 1.03), n=4 | Reference |
| **Maternal composite outcome** | **Pre-alpha** | Reference | 1.14 (0.85, 1.53), n=4 | 1.73 (0.92, 3.26), n=1 | | 0.82 (0.52, 1.32), n=3 | n=0 | | 1.12 (0.55, 2.26), n=1 | 0.61 (0.26, 1.41), n=3 | 1.20 (0.93, 1.55), n=5 |
|  | **Alpha** | 0.87 (0.65, 1.17), n=4 | Reference | 1.00 (0.49, 2.05), n=1 | | 0.92 (0.63, 1.34), n=3 | n=0 | | 0.90 (0.41, 1.98), n=1 | 0.49 (0.25, 0.96), n=3 | 1.07 (0.76, 1.52), n=5 |
|  | **Beta** | 0.58 (0.31, 1.09), n=1 | 1.00 (0.49, 2.05), n=1 | Reference | | n=0 | n=0 | | n=0 | 0.50 (0.29, 0.88), n=1 | 0.90 (0.35, 2.32), n=1 |
|  | **Delta** | 1.21 (0.76, 1.94), n=3 | 1.08 (0.74, 1.58), n=3 | n=0 | | Reference | n=0 | | 0.81 (0.39, 1.70), n=1 | 0.72 (0.51, 1.03), n=2 | 1.23 (0.80, 1.89), n=3 |
|  | **Epsilon** | n=0 | n=0 | n=0 | | n=0 | Reference | | n=0 | n=0 | n=0 |
|  | **Eta** | 0.89 (0.44, 1.80), n=1 | 1.11 (0.50, 2.44), n=1 | n=0 | | 1.23 (0.59, 2.58), n=1 | n=0 | | Reference | 1.00 (0.50, 2.02), n=1 | 1.09 (0.53, 2.21), n=1 |
|  | **Omicron** | 1.65 (0.71, 3.83), n=3 | 2.02 (1.04, 3.93), n=3 | 2.00 (1.14, 3.51), n=1 | | 1.39 (0.98, 1.97), n=2 | n=0 | | 1.00 (0.49, 2.01), n=1 | Reference | 1.85 (0.92, 3.71), n=3 |
|  | **Mixed** | 0.83 (0.65, 1.07), n=5 | 0.93 (0.66, 1.32), n=5 | 1.11 (0.43, 2.86), n=1 | | 0.81 (0.53, 1.24), n=3 | n=0 | | 0.92 (0.45, 1.87), n=1 | 0.54 (0.27, 1.09), n=3 | Reference |

HDP: Hypertensive disorders of pregnancy

C. **Fetal and Neonatal Morbidity & Mortality**

| **Outcome** | **Reference Strain** | **Pre-alpha** | **Alpha** | **Beta** | **Delta** | **Epsilon** | **Eta** | **Omicron** | **Mixed** |
| --- | --- | --- | --- | --- | --- | --- | --- | --- | --- |
| **Stillbirth** | **Pre-alpha** | Reference | 2.07 (0.27, 15.57), n=2 | n=0 | 3.79 (1.03, 13.98), n=3 | n=0 | n=0 | 1.30 (0.27, 6.21), n=3 | 6.11 (1.02, 36.68), n=2 |
|  | **Alpha** | 0.48 (0.06, 3.65), n=2 | Reference | n=0 | 1.89 (0.38, 9.49), n=3 | n=0 | n=0 | 0.96 (0.16, 5.76), n=3 | 2.88 (0.30, 27.21), n=1 |
|  | **Beta** | n=0 | n=0 | Reference | n=0 | n=0 | n=0 | n=0 | n=0 |
|  | **Delta** | 0.26 (0.07, 0.97), n=3 | 0.53 (0.11, 2.66), n=3 | n=0 | Reference | n=0 | n=0 | 0.35 (0.07, 1.69), n=3 | 1.15 (0.29, 4.54), n=3 |
|  | **Eta** | n=0 | n=0 | n=0 | n=0 | n=0 | Reference | n=0 | n=0 |
|  | **Omicron** | 0.77 (0.16, 3.69), n=3 | 1.05 (0.17, 6.29), n=3 | n=0 | 2.88 (0.59, 14.05), n=3 | n=0 | n=0 | Reference | 3.50 (0.61, 20.05), n=3 |
|  | **Mixed** | 0.16 (0.03, 0.98), n=2 | 0.35 (0.04, 3.28), n=1 | n=0 | 0.87 (0.22, 3.45), n=3 | n=0 | n=0 | 0.29 (0.05, 1.63), n=3 | Reference |
| **Perinatal death** | **Pre-alpha** | Reference | 1.70 (0.25, 11.55), n=2 | n=0 | 4.00 (0.93, 17.18), n=3 | n=0 | n=0 | 2.92 (0.59, 14.43), n=2 | 7.23 (1.53, 34.06), n=2 |
|  | **Alpha** | 0.59 (0.09, 3.98), n=2 | Reference | n=0 | 1.68 (0.26, 10.90), n=2 | n=0 | n=0 | 1.55 (0.18, 13.16), n=2 | 2.88 (0.30, 27.21), n=1 |
|  | **Beta** | n=0 | n=0 | Reference | n=0 | n=0 | n=0 | n=0 | n=0 |
|  | **Delta** | 0.25 (0.06, 1.07), n=3 | 0.60 (0.09, 3.86), n=2 | n=0 | Reference | n=0 | n=0 | 0.68 (0.08, 5.94), n=2 | 1.30 (0.28, 5.98), n=2 |
|  | **Omicron** | 0.34 (0.07, 1.69), n=2 | 0.65 (0.08, 5.51), n=2 | n=0 | 1.48 (0.17, 12.92), n=2 | n=0 | n=0 | Reference | 3.51 (0.60, 20.59), n=2 |
|  | **Mixed** | 0.14 (0.03, 0.65), n=2 | 0.35 (0.04, 3.28), n=1 | n=0 | 0.77 (0.17, 3.52), n=2 | n=0 | n=0 | 0.29 (0.05, 1.68), n=2 | Reference |
| **Early neonatal death** | **Pre-alpha** | Reference | 1.12 (0.12, 10.73), n=2 | n=0 | 1.86 (0.29, 11.74), n=3 | n=0 | n=0 | 2.42 (0.39, 14.77), n=2 | 2.89 (0.30, 27.47), n=2 |
|  | **Alpha** | 0.89 (0.09, 8.54), n=2 | Reference | n=0 | 0.74 (0.03, 17.77), n=1 | n=0 | n=0 | 1.99 (0.22, 17.89), n=2 | n=0 |
|  | **Beta** | n=0 | n=0 | Reference | n=0 | n=0 | n=0 | n=0 | n=0 |
|  | **Delta** | 0.54 (0.09, 3.40), n=3 | 1.36 (0.06, 32.73), n=1 | n=0 | Reference | n=0 | n=0 | 1.41 (0.16, 12.71), n=2 | 1.73 (0.07, 41.49), n=1 |
|  | **Omicron** | 0.41 (0.07, 2.53), n=2 | 0.50 (0.06, 4.53), n=2 | n=0 | 0.71 (0.08, 6.36), n=2 | n=0 | n=0 | Reference | 1.37 (0.15, 12.19), n=2 |
|  | **Mixed** | 0.35 (0.04, 3.29), n=2 | n=0 | n=0 | 0.58 (0.02, 13.91), n=1 | n=0 | n=0 | 0.73 (0.08, 6.54), n=2 | Reference |
| **Neonatal death** | **Pre-alpha** | Reference | 1.12 (0.12, 10.73), n=2 | n=0 | 2.94 (0.60, 14.49), n=4 | n=0 | n=0 | 2.42 (0.39, 14.77), n=2 | 2.89 (0.30, 27.47), n=2 |
|  | **Alpha** | 0.89 (0.09, 8.54), n=2 | Reference | n=0 | 1.23 (0.13, 11.63), n=2 | n=0 | n=0 | 1.99 (0.22, 17.89), n=2 | n=0 |
|  | **Beta** | n=0 | n=0 | Reference | n=0 | n=0 | n=0 | n=0 | n=0 |
|  | **Delta** | 0.34 (0.07, 1.68), n=4 | 0.81 (0.09, 7.63), n=2 | n=0 | Reference | n=0 | 0.97 (0.04, 22.47), n=1 | 0.61 (0.10, 3.70), n=3 | 0.51 (0.05, 4.80), n=2 |
|  | **Eta** | n=0 | n=0 | n=0 | 1.03 (0.04, 23.67), n=1 | n=0 | Reference | n=0 | n=0 |
|  | **Omicron** | 0.41 (0.07, 2.53), n=2 | 0.50 (0.06, 4.53), n=2 | n=0 | 1.65 (0.27, 10.04), n=3 | n=0 | n=0 | Reference | 1.37 (0.15, 12.19), n=2 |
|  | **Mixed** | 0.35 (0.04, 3.29), n=2 | n=0 | n=0 | 1.97 (0.21, 18.66), n=2 | n=0 | n=0 | 0.73 (0.08, 6.54), n=2 | Reference |
| **NICU** | **Pre-alpha** | Reference | 0.72 (0.09, 5.55), n=1 | n=0 | 0.49 (0.06, 3.79), n=1 | n=0 | 1.51 (0.21, 11.09), n=1 | 1.19 (0.46, 3.07), n=1 | 0.86 (0.27, 2.78), n=1 |
|  | **Alpha** | 1.38 (0.18, 10.56), n=1 | Reference | n=0 | 0.68 (0.04, 10.31), n=1 | n=0 | 2.08 (0.14, 30.54), n=1 | 1.64 (0.21, 12.53), n=1 | 1.19 (0.14, 10.17), n=1 |
|  | **Delta** | 2.04 (0.26, 15.81), n=1 | 1.48 (0.10, 22.58), n=1 | n=0 | Reference | n=0 | 3.08 (0.21, 45.62), n=1 | 2.43 (0.31, 18.77), n=1 | 1.76 (0.20, 15.23), n=1 |
|  | **Eta** | 0.66 (0.09, 4.86), n=1 | 0.48 (0.03, 7.04), n=1 | n=0 | 0.32 (0.02, 4.80), n=1 | n=0 | Reference | 0.79 (0.11, 5.77), n=1 | 0.57 (0.07, 4.70), n=1 |
|  | **Omicron** | 0.84 (0.33, 2.18), n=1 | 0.61 (0.08, 4.66), n=1 | n=0 | 0.41 (0.05, 3.19), n=1 | n=0 | 1.27 (0.17, 9.32), n=1 | Reference | 0.73 (0.23, 2.33), n=1 |
|  | **Mixed** | 1.16 (0.36, 3.73), n=1 | 0.84 (0.10, 7.18), n=1 | n=0 | 0.57 (0.07, 4.91), n=1 | n=0 | 1.75 (0.21, 14.38), n=1 | 1.38 (0.43, 4.43), n=1 | Reference |
| **Fetal composite outcome** | **Pre-alpha** | Reference | 0.98 (0.55, 1.74), n=5 | 0.62 (0.05, 8.41), n=1 | 0.79 (0.46, 1.35), n=3 | n=0 | 0.50 (0.07, 3.41), n=1 | 0.42 (0.15, 1.22), n=3 | 1.64 (0.87, 3.08), n=5 |
|  | **Alpha** | 1.02 (0.58, 1.82), n=5 | Reference | n=0 | 0.96 (0.59, 1.56), n=3 | n=0 | 0.54 (0.07, 4.34), n=1 | 0.50 (0.17, 1.42), n=3 | 1.33 (0.71, 2.47), n=5 |
|  | **Beta** | 1.61 (0.12, 21.71), n=1 | n=0 | Reference | n=0 | n=0 | n=0 | 0.97 (0.07, 12.68), n=1 | 4.50 (0.32, 63.94), n=1 |
|  | **Delta** | 1.26 (0.74, 2.15), n=3 | 1.05 (0.64, 1.70), n=3 | n=0 | Reference | n=0 | 0.62 (0.08, 4.77), n=1 | 0.53 (0.17, 1.69), n=2 | 1.95 (1.24, 3.07), n=3 |
|  | **Epsilon** | n=0 | n=0 | n=0 | n=0 | Reference | n=0 | n=0 | n=0 |
|  | **Eta** | 1.99 (0.29, 13.44), n=1 | 1.85 (0.23, 14.80), n=1 | n=0 | 1.62 (0.21, 12.55), n=1 | n=0 | Reference | 2.07 (0.30, 14.04), n=1 | 1.43 (0.20, 10.19), n=1 |
|  | **Omicron** | 2.36 (0.82, 6.83), n=3 | 2.02 (0.70, 5.79), n=3 | 1.03 (0.08, 13.57), n=1 | 1.89 (0.59, 6.03), n=2 | n=0 | 0.48 (0.07, 3.29), n=1 | Reference | 2.81 (0.86, 9.23), n=3 |
|  | **Mixed** | 0.61 (0.32, 1.15), n=5 | 0.75 (0.40, 1.40), n=5 | 0.22 (0.02, 3.16), n=1 | 0.51 (0.33, 0.81), n=3 | n=0 | 0.70 (0.10, 4.99), n=1 | 0.36 (0.11, 1.17), n=3 | Reference |

NICU: Neonatal intensive care unit

D. **Adverse Birth Outcomes**

| **Outcome** | **Reference Strain** | **Pre-alpha** | **Alpha** | **Beta** | **Delta** | **Epsilon** | **Eta** | **Omicron** | **Mixed** |
| --- | --- | --- | --- | --- | --- | --- | --- | --- | --- |
| **vLBW** | **Pre-alpha** | Reference | 1.21 (0.39, 3.72), n=3 | n=0 | 4.10 (1.75, 9.60), n=4 | n=0 | 3.74 (0.16, 87.35), n=1 | 1.32 (0.59, 2.95), n=4 | 1.81 (0.52, 6.27), n=4 |
|  | **Alpha** | 0.83 (0.27, 2.56), n=3 | Reference | n=0 | 2.07 (0.54, 7.89), n=4 | n=0 | n=0 | 0.83 (0.29, 2.40), n=4 | 2.00 (0.44, 9.15), n=3 |
|  | **Beta** | n=0 | n=0 | Reference | n=0 | n=0 | n=0 | 0.10 (0.01, 1.98), n=1 | n=0 |
|  | **Delta** | 0.24 (0.10, 0.57), n=4 | 0.48 (0.13, 1.85), n=4 | n=0 | Reference | n=0 | 0.97 (0.04, 22.47), n=1 | 0.44 (0.13, 1.50), n=3 | 0.76 (0.13, 4.45), n=4 |
|  | **Eta** | 0.27 (0.01, 6.23), n=1 | n=0 | n=0 | 1.03 (0.04, 23.67), n=1 | n=0 | Reference | 0.74 (0.04, 13.55), n=1 | n=0 |
|  | **Omicron** | 0.76 (0.34, 1.71), n=4 | 1.20 (0.42, 3.46), n=4 | 10.00 (0.50, 198.21), n=1 | 2.26 (0.66, 7.67), n=3 | n=0 | 1.35 (0.07, 24.75), n=1 | Reference | 1.49 (0.44, 5.10), n=4 |
|  | **Mixed** | 0.55 (0.16, 1.92), n=4 | 0.50 (0.11, 2.28), n=3 | n=0 | 1.31 (0.22, 7.68), n=4 | n=0 | n=0 | 0.67 (0.20, 2.29), n=4 | Reference |
| **LBW** | **Pre-alpha** | Reference | 1.13 (0.77, 1.67), n=5 | 0.85 (0.06, 11.88), n=1 | 1.40 (0.92, 2.14), n=4 | n=0 | 1.21 (0.17, 8.66), n=1 | 1.02 (0.76, 1.37), n=4 | 1.61 (0.79, 3.31), n=5 |
|  | **Alpha** | 0.88 (0.60, 1.31), n=5 | Reference | n=0 | 1.23 (0.59, 2.56), n=4 | n=0 | 2.08 (0.14, 30.54), n=1 | 0.91 (0.59, 1.41), n=4 | 1.43 (0.76, 2.67), n=5 |
|  | **Beta** | 1.18 (0.08, 16.50), n=1 | n=0 | Reference | n=0 | n=0 | n=0 | 0.30 (0.02, 4.40), n=1 | 4.50 (0.32, 63.94), n=1 |
|  | **Delta** | 0.71 (0.47, 1.09), n=4 | 0.82 (0.39, 1.70), n=4 | n=0 | Reference | n=0 | 1.03 (0.12, 8.98), n=1 | 0.90 (0.54, 1.51), n=3 | 1.01 (0.49, 2.08), n=4 |
|  | **Eta** | 0.83 (0.12, 5.93), n=1 | 0.48 (0.03, 7.04), n=1 | n=0 | 0.97 (0.11, 8.50), n=1 | n=0 | Reference | 1.67 (0.24, 11.49), n=1 | 0.57 (0.07, 4.70), n=1 |
|  | **Omicron** | 0.98 (0.73, 1.32), n=4 | 1.10 (0.71, 1.70), n=4 | 3.33 (0.23, 48.90), n=1 | 1.11 (0.66, 1.87), n=3 | n=0 | 0.60 (0.09, 4.11), n=1 | Reference | 1.44 (0.38, 5.49), n=4 |
|  | **Mixed** | 0.62 (0.30, 1.27), n=5 | 0.70 (0.37, 1.31), n=5 | 0.22 (0.02, 3.16), n=1 | 0.99 (0.48, 2.05), n=4 | n=0 | 1.75 (0.21, 14.38), n=1 | 0.69 (0.18, 2.65), n=4 | Reference |
| **eSGA** | **Pre-alpha** | Reference | 1.54 (0.72, 3.30), n=4 | 3.11 (0.16, 60.70), n=1 | 1.55 (0.67, 3.58), n=3 | n=0 | n=0 | 0.39 (0.14, 1.08), n=3 | 2.79 (0.75, 10.37), n=4 |
|  | **Alpha** | 0.65 (0.30, 1.39), n=4 | Reference | n=0 | 0.94 (0.30, 2.91), n=3 | n=0 | n=0 | 0.32 (0.10, 1.02), n=1 | 1.76 (0.39, 7.81), n=3 |
|  | **Beta** | 0.32 (0.02, 6.27), n=1 | n=0 | Reference | n=0 | n=0 | n=0 | n=0 | 4.50 (0.32, 63.94), n=1 |
|  | **Delta** | 0.64 (0.28, 1.49), n=3 | 1.06 (0.34, 3.29), n=3 | n=0 | Reference | n=0 | n=0 | 0.43 (0.10, 1.89), n=2 | 0.87 (0.25, 2.96), n=3 |
|  | **Omicron** | 2.55 (0.93, 6.99), n=3 | 3.12 (0.98, 9.96), n=1 | n=0 | 2.30 (0.53, 9.99), n=2 | n=0 | n=0 | Reference | 11.90 (0.41, 346.81), n=2 |
|  | **Mixed** | 0.36 (0.10, 1.33), n=4 | 0.57 (0.13, 2.53), n=3 | 0.22 (0.02, 3.16), n=1 | 1.15 (0.34, 3.93), n=3 | n=0 | n=0 | 0.08 (0.00, 2.45), n=2 | Reference |
| **SGA** | **Pre-alpha** | Reference | 1.43 (0.73, 2.82), n=4 | 1.87 (0.11, 30.49), n=1 | 1.65 (0.89, 3.08), n=3 | n=0 | n=0 | 0.60 (0.25, 1.44), n=3 | 3.49 (1.70, 7.16), n=4 |
|  | **Alpha** | 0.70 (0.35, 1.38), n=4 | Reference | n=0 | 1.12 (0.46, 2.71), n=3 | n=0 | n=0 | 0.34 (0.12, 1.00), n=2 | 2.36 (0.82, 6.81), n=3 |
|  | **Beta** | 0.54 (0.03, 8.75), n=1 | n=0 | Reference | n=0 | n=0 | n=0 | 0.50 (0.04, 6.88), n=1 | 4.50 (0.32, 63.94), n=1 |
|  | **Delta** | 0.60 (0.33, 1.12), n=3 | 0.89 (0.37, 2.15), n=3 | n=0 | Reference | n=0 | n=0 | 0.43 (0.10, 1.89), n=2 | 1.53 (0.74, 3.17), n=3 |
|  | **Omicron** | 1.66 (0.69, 3.96), n=3 | 2.90 (1.00, 8.39), n=2 | 2.00 (0.15, 27.51), n=1 | 2.30 (0.53, 9.99), n=2 | n=0 | n=0 | Reference | 6.99 (2.60, 18.81), n=2 |
|  | **Mixed** | 0.29 (0.14, 0.59), n=4 | 0.42 (0.15, 1.23), n=3 | 0.22 (0.02, 3.16), n=1 | 0.65 (0.32, 1.35), n=3 | n=0 | n=0 | 0.14 (0.05, 0.38), n=2 | Reference |
| **vPTB** | **Pre-alpha** | Reference | 1.52 (0.81, 2.87), n=5 | 3.11 (0.16, 60.70), n=1 | 3.34 (1.98, 5.65), n=4 | n=0 | 6.04 (0.59, 61.92), n=1 | 1.02 (0.58, 1.78), n=4 | 1.46 (0.64, 3.33), n=5 |
|  | **Alpha** | 0.66 (0.35, 1.24), n=5 | Reference | n=0 | 1.90 (0.93, 3.88), n=4 | n=0 | 2.17 (0.15, 31.79), n=1 | 0.63 (0.32, 1.26), n=4 | 0.88 (0.32, 2.41), n=4 |
|  | **Beta** | 0.32 (0.02, 6.27), n=1 | n=0 | Reference | n=0 | n=0 | n=0 | 0.17 (0.01, 2.77), n=1 | n=0 |
|  | **Delta** | 0.30 (0.18, 0.51), n=4 | 0.53 (0.26, 1.07), n=4 | n=0 | Reference | n=0 | 3.08 (0.21, 45.62), n=1 | 0.35 (0.16, 0.77), n=3 | 0.46 (0.18, 1.19), n=4 |
|  | **Eta** | 0.17 (0.02, 1.70), n=1 | 0.46 (0.03, 6.77), n=1 | n=0 | 0.32 (0.02, 4.80), n=1 | n=0 | Reference | 0.49 (0.06, 3.87), n=1 | 0.29 (0.03, 2.92), n=1 |
|  | **Omicron** | 0.98 (0.56, 1.72), n=4 | 1.59 (0.79, 3.17), n=4 | 6.00 (0.36, 99.66), n=1 | 2.83 (1.30, 6.15), n=3 | n=0 | 2.03 (0.26, 16.01), n=1 | Reference | 1.17 (0.49, 2.78), n=4 |
|  | **Mixed** | 0.69 (0.30, 1.57), n=5 | 1.14 (0.42, 3.12), n=4 | n=0 | 2.16 (0.84, 5.55), n=4 | n=0 | 3.50 (0.34, 35.72), n=1 | 0.85 (0.36, 2.02), n=4 | Reference |
| **vPTB (COVID-19 onset <34w)** | **Pre-alpha** | Reference | n=0 | n=0 | n=0 | n=0 | n=0 | n=0 | n=0 |
|  | **Alpha** | n=0 | Reference | n=0 | n=0 | n=0 | n=0 | n=0 | n=0 |
|  | **Delta** | n=0 | n=0 | n=0 | Reference | n=0 | n=0 | n=0 | n=0 |
|  | **Omicron** | n=0 | n=0 | n=0 | n=0 | n=0 | n=0 | Reference | n=0 |
|  | **Mixed** | n=0 | n=0 | n=0 | n=0 | n=0 | n=0 | n=0 | Reference |
| **PTB** | **Pre-alpha** | Reference | 0.86 (0.56, 1.31), n=5 | 1.04 (0.07, 14.95), n=1 | 1.52 (0.87, 2.67), n=4 | n=0 | 1.51 (0.21, 11.09), n=1 | 1.06 (0.79, 1.44), n=4 | 1.45 (0.82, 2.57), n=5 |
|  | **Alpha** | 1.17 (0.76, 1.78), n=5 | Reference | n=0 | 1.58 (0.93, 2.69), n=4 | n=0 | 2.17 (0.15, 31.79), n=1 | 1.13 (0.71, 1.79), n=4 | 1.24 (0.62, 2.51), n=4 |
|  | **Beta** | 0.96 (0.07, 13.90), n=1 | n=0 | Reference | n=0 | n=0 | n=0 | 0.63 (0.05, 8.53), n=1 | n=0 |
|  | **Delta** | 0.66 (0.37, 1.15), n=4 | 0.63 (0.37, 1.08), n=4 | n=0 | Reference | n=0 | 1.54 (0.15, 15.54), n=1 | 0.74 (0.48, 1.14), n=3 | 0.98 (0.45, 2.11), n=4 |
|  | **Eta** | 0.66 (0.09, 4.86), n=1 | 0.46 (0.03, 6.77), n=1 | n=0 | 0.65 (0.06, 6.54), n=1 | n=0 | Reference | 1.08 (0.15, 7.68), n=1 | 1.14 (0.16, 8.35), n=1 |
|  | **Omicron** | 0.94 (0.70, 1.27), n=4 | 0.88 (0.56, 1.40), n=4 | 1.58 (0.12, 21.28), n=1 | 1.35 (0.88, 2.08), n=3 | n=0 | 0.92 (0.13, 6.56), n=1 | Reference | 0.90 (0.53, 1.52), n=4 |
|  | **Mixed** | 0.69 (0.39, 1.22), n=5 | 0.80 (0.40, 1.62), n=4 | n=0 | 1.02 (0.47, 2.21), n=4 | n=0 | 0.88 (0.12, 6.39), n=1 | 1.11 (0.66, 1.89), n=4 | Reference |
| **PTB (COVID-19 onset <37w)** | **Pre-alpha** | Reference | 0.86 (0.55, 1.36), n=2 | n=0 | 1.59 (0.83, 3.05), n=2 | n=0 | n=0 | 1.09 (0.67, 1.77), n=2 | 0.82 (0.40, 1.65), n=2 |
|  | **Alpha** | 1.16 (0.74, 1.83), n=2 | Reference | n=0 | 1.64 (0.92, 2.92), n=2 | n=0 | n=0 | 1.13 (0.69, 1.86), n=2 | 0.85 (0.37, 1.97), n=2 |
|  | **Delta** | 0.63 (0.33, 1.20), n=2 | 0.61 (0.34, 1.09), n=2 | n=0 | Reference | n=0 | n=0 | 0.68 (0.43, 1.08), n=2 | 0.44 (0.20, 0.94), n=2 |
|  | **Omicron** | 0.92 (0.56, 1.50), n=2 | 0.88 (0.54, 1.46), n=2 | n=0 | 1.46 (0.92, 2.31), n=2 | n=0 | n=0 | Reference | 0.65 (0.32, 1.35), n=2 |
|  | **Mixed** | 1.23 (0.61, 2.47), n=2 | 1.17 (0.51, 2.70), n=2 | n=0 | 2.28 (1.07, 4.88), n=2 | n=0 | n=0 | 1.53 (0.74, 3.14), n=2 | Reference |

vLBW: Very low birthweight; LBW: Low birthweight; eSGA: Extremely small-for-gestational age; SGA: Small-for-gestational age; vPTB: Very Preterm birth; PTB: Preterm birth

**Table S2. Random effect relative risk (RR) estimates with 95% confidence intervals (CI) and number of studies included in each analysis (dominant strain threshold ≥70%)**

A. **COVID-19 Severity Outcomes**

| **Outcome** | **Reference Strain** | **Pre-alpha** | **Alpha** | **Delta** | **Eta** | **Omicron** | **Mixed** |
| --- | --- | --- | --- | --- | --- | --- | --- |
| **Hospitalization** | **Pre-alpha** | Reference | 1.42 (0.75, 2.68), n=4 | 1.05 (0.42, 2.63), n=2 | 1.21 (0.17, 8.66), n=1 | 0.99 (0.87, 1.11), n=2 | 1.17 (1.02, 1.35), n=5 |
|  | **Alpha** | 0.71 (0.37, 1.34), n=4 | Reference | 0.54 (0.22, 1.31), n=2 | 0.35 (0.05, 2.57), n=1 | 0.40 (0.08, 1.99), n=2 | 0.79 (0.46, 1.35), n=4 |
|  | **Delta** | 0.95 (0.38, 2.38), n=2 | 1.85 (0.76, 4.48), n=2 | Reference | 0.62 (0.08, 4.77), n=1 | 0.18 (0.05, 0.73), n=1 | 0.66 (0.25, 1.72), n=2 |
|  | **Eta** | 0.83 (0.12, 5.93), n=1 | 2.88 (0.39, 21.32), n=1 | 1.62 (0.21, 12.55), n=1 | Reference | 0.30 (0.03, 2.62), n=1 | 0.99 (0.13, 7.35), n=1 |
|  | **Omicron** | 1.01 (0.90, 1.15), n=2 | 2.53 (0.50, 12.67), n=2 | 5.50 (1.38, 21.92), n=1 | 3.39 (0.38, 30.10), n=1 | Reference | 1.05 (0.73, 1.50), n=2 |
|  | **Mixed** | 0.85 (0.74, 0.98), n=5 | 1.27 (0.74, 2.18), n=4 | 1.52 (0.58, 3.96), n=2 | 1.01 (0.14, 7.52), n=1 | 0.95 (0.66, 1.37), n=2 | Reference |
| **ICU admission** | **Pre-alpha** | Reference | 1.36 (0.71, 2.60), n=4 | 2.13 (0.95, 4.77), n=3 | 12.08 (0.81, 181.34), n=1 | 0.67 (0.22, 2.04), n=4 | 1.12 (0.73, 1.73), n=5 |
|  | **Alpha** | 0.74 (0.38, 1.41), n=4 | Reference | 1.37 (0.62, 3.02), n=3 | 6.00 (0.26, 137.33), n=1 | 0.40 (0.15, 1.08), n=4 | 0.92 (0.39, 2.16), n=3 |
|  | **Delta** | 0.47 (0.21, 1.05), n=3 | 0.73 (0.33, 1.61), n=3 | Reference | 3.08 (0.21, 45.62), n=1 | 0.34 (0.15, 0.73), n=3 | 0.68 (0.30, 1.55), n=3 |
|  | **Eta** | 0.08 (0.01, 1.24), n=1 | 0.17 (0.01, 3.81), n=1 | 0.32 (0.02, 4.80), n=1 | Reference | 0.10 (0.01, 1.47), n=1 | 0.42 (0.05, 3.75), n=1 |
|  | **Omicron** | 1.49 (0.49, 4.55), n=4 | 2.51 (0.92, 6.82), n=4 | 2.98 (1.37, 6.47), n=3 | 10.17 (0.68, 152.44), n=1 | Reference | 2.37 (0.97, 5.75), n=4 |
|  | **Mixed** | 0.89 (0.58, 1.38), n=5 | 1.09 (0.46, 2.56), n=3 | 1.46 (0.65, 3.31), n=3 | 2.36 (0.27, 20.91), n=1 | 0.42 (0.17, 1.03), n=4 | Reference |
| **Critical care** | **Pre-alpha** | Reference | 1.26 (0.66, 2.42), n=4 | 1.88 (0.95, 3.74), n=4 | 6.04 (0.59, 61.92), n=1 | 0.70 (0.20, 2.46), n=3 | 0.98 (0.64, 1.50), n=5 |
|  | **Alpha** | 0.79 (0.41, 1.52), n=4 | Reference | 1.48 (0.69, 3.20), n=3 | 6.00 (0.26, 137.33), n=1 | 0.49 (0.16, 1.50), n=3 | 0.92 (0.39, 2.16), n=3 |
|  | **Delta** | 0.53 (0.27, 1.05), n=4 | 0.67 (0.31, 1.46), n=3 | Reference | 3.08 (0.21, 45.62), n=1 | 0.34 (0.16, 0.73), n=3 | 0.64 (0.28, 1.42), n=3 |
|  | **Eta** | 0.17 (0.02, 1.70), n=1 | 0.17 (0.01, 3.81), n=1 | 0.32 (0.02, 4.80), n=1 | Reference | 0.10 (0.01, 1.47), n=1 | 0.42 (0.05, 3.75), n=1 |
|  | **Omicron** | 1.42 (0.41, 4.95), n=3 | 2.03 (0.67, 6.15), n=3 | 2.94 (1.38, 6.29), n=3 | 10.17 (0.68, 152.44), n=1 | Reference | 2.05 (0.77, 5.43), n=3 |
|  | **Mixed** | 1.02 (0.67, 1.57), n=5 | 1.09 (0.46, 2.56), n=3 | 1.57 (0.71, 3.51), n=3 | 2.36 (0.27, 20.91), n=1 | 0.49 (0.18, 1.30), n=3 | Reference |
| **Ventilation** | **Pre-alpha** | Reference | 2.26 (0.93, 5.46), n=5 | 2.79 (1.16, 6.75), n=4 | 6.04 (0.59, 61.92), n=1 | 0.73 (0.27, 1.94), n=4 | 1.10 (0.40, 3.05), n=6 |
|  | **Alpha** | 0.44 (0.18, 1.07), n=5 | Reference | 1.62 (0.61, 4.25), n=2 | 6.00 (0.26, 137.33), n=1 | 0.30 (0.11, 0.83), n=4 | 0.84 (0.29, 2.45), n=3 |
|  | **Delta** | 0.36 (0.15, 0.86), n=4 | 0.62 (0.24, 1.63), n=2 | Reference | 8.77 (0.38, 202.21), n=1 | 0.23 (0.08, 0.65), n=3 | 0.57 (0.20, 1.57), n=3 |
|  | **Eta** | 0.17 (0.02, 1.70), n=1 | 0.17 (0.01, 3.81), n=1 | 0.11 (0.00, 2.63), n=1 | Reference | 0.10 (0.01, 1.47), n=1 | 0.14 (0.01, 2.11), n=1 |
|  | **Omicron** | 1.38 (0.52, 3.67), n=4 | 3.33 (1.21, 9.18), n=4 | 4.31 (1.55, 12.01), n=3 | 10.17 (0.68, 152.44), n=1 | Reference | 2.72 (0.95, 7.79), n=4 |
|  | **Mixed** | 0.91 (0.33, 2.50), n=6 | 1.19 (0.41, 3.48), n=3 | 1.76 (0.64, 4.88), n=3 | 7.08 (0.47, 105.94), n=1 | 0.37 (0.13, 1.05), n=4 | Reference |
| **Pneumonia** | **Pre-alpha** | Reference | 1.13 (0.55, 2.31), n=4 | 0.70 (0.27, 1.82), n=4 | 0.76 (0.11, 5.22), n=1 | 0.30 (0.10, 0.87), n=3 | 1.34 (1.07, 1.68), n=5 |
|  | **Alpha** | 0.89 (0.43, 1.81), n=4 | Reference | 0.60 (0.23, 1.58), n=4 | 0.52 (0.07, 4.17), n=1 | 0.28 (0.05, 1.56), n=3 | 0.82 (0.34, 2.00), n=3 |
|  | **Delta** | 1.43 (0.55, 3.70), n=4 | 1.66 (0.63, 4.35), n=4 | Reference | 0.77 (0.10, 6.25), n=1 | 0.20 (0.05, 0.71), n=3 | 1.21 (0.53, 2.74), n=4 |
|  | **Eta** | 1.32 (0.19, 9.15), n=1 | 1.92 (0.24, 15.37), n=1 | 1.30 (0.16, 10.51), n=1 | Reference | 0.10 (0.01, 1.47), n=1 | 1.27 (0.18, 9.16), n=1 |
|  | **Omicron** | 3.39 (1.15, 10.02), n=3 | 3.52 (0.64, 19.29), n=3 | 5.07 (1.41, 18.25), n=3 | 10.17 (0.68, 152.44), n=1 | Reference | 6.41 (1.97, 20.90), n=3 |
|  | **Mixed** | 0.74 (0.59, 0.93), n=5 | 1.22 (0.50, 2.96), n=3 | 0.83 (0.37, 1.88), n=4 | 0.79 (0.11, 5.68), n=1 | 0.16 (0.05, 0.51), n=3 | Reference |

*Hospitalization was categorized as follows:

Martinez-Portilla, 2021: Clearly defined hospitalization for COVID-19 | Gill, Fernandez Buhigas, 2021: Clearly defined hospitalization for COVID-19 | Bevilacqua, Laurita Longo, 2020: Hospitalization for COVID-19 included those for L&D as well as pregnancy-related preterm labor/births. | Akelo, Tippett Barr 2021: Clearly defined hospitalization for COVID-19 | Poon 2021: Hospitalization for COVD-19 included any hospital admission

B. **Maternal Morbidity & Mortality**

| **Outcome** | **Reference Strain** | **Pre-alpha** | **Alpha** | **Delta** | **Eta** | **Omicron** | **Mixed** |
| --- | --- | --- | --- | --- | --- | --- | --- |
| **Pregnancy-related death** | **Pre-alpha** | Reference | 1.29 (0.12, 14.11), n=1 | 0.68 (0.03, 13.96), n=1 | n=0 | 0.26 (0.01, 5.44), n=1 | 0.32 (0.09, 1.15), n=2 |
|  | **Alpha** | 0.77 (0.07, 8.46), n=1 | Reference | 0.44 (0.02, 10.64), n=1 | n=0 | 0.17 (0.01, 4.15), n=1 | 0.42 (0.02, 10.20), n=1 |
|  | **Delta** | 1.48 (0.07, 30.54), n=1 | 2.28 (0.09, 55.35), n=1 | Reference | n=0 | n=0 | n=0 |
|  | **Eta** | n=0 | n=0 | n=0 | Reference | n=0 | n=0 |
|  | **Omicron** | 3.81 (0.18, 78.99), n=1 | 5.88 (0.24, 143.16), n=1 | n=0 | n=0 | Reference | n=0 |
|  | **Mixed** | 3.09 (0.87, 10.95), n=2 | 2.38 (0.10, 57.77), n=1 | n=0 | n=0 | n=0 | Reference |
| **Placental abruption** | **Pre-alpha** | Reference | 2.16 (0.39, 11.90), n=3 | 0.44 (0.05, 4.22), n=2 | n=0 | 0.35 (0.04, 2.83), n=2 | 2.78 (0.54, 14.34), n=3 |
|  | **Alpha** | 0.46 (0.08, 2.55), n=3 | Reference | 0.06 (0.00, 1.35), n=1 | n=0 | 0.10 (0.01, 1.98), n=1 | 1.63 (0.20, 13.05), n=2 |
|  | **Delta** | 2.27 (0.24, 21.69), n=2 | 17.93 (0.74, 433.21), n=1 | Reference | n=0 | n=0 | 6.74 (0.71, 64.16), n=2 |
|  | **Omicron** | 2.82 (0.35, 22.59), n=2 | 10.00 (0.50, 198.21), n=1 | n=0 | n=0 | Reference | 7.46 (0.83, 67.37), n=2 |
|  | **Mixed** | 0.36 (0.07, 1.85), n=3 | 0.61 (0.08, 4.90), n=2 | 0.15 (0.02, 1.41), n=2 | n=0 | 0.13 (0.01, 1.21), n=2 | Reference |
| **Preterm labor** | **Pre-alpha** | Reference | 0.69 (0.27, 1.77), n=4 | 1.53 (0.76, 3.11), n=3 | 2.25 (0.11, 44.35), n=1 | 0.83 (0.44, 1.59), n=3 | 1.45 (0.70, 3.03), n=4 |
|  | **Alpha** | 1.45 (0.56, 3.71), n=4 | Reference | 2.14 (0.70, 6.49), n=3 | n=0 | 1.14 (0.38, 3.41), n=3 | 2.06 (0.67, 6.36), n=3 |
|  | **Delta** | 0.65 (0.32, 1.32), n=3 | 0.47 (0.15, 1.42), n=3 | Reference | 0.97 (0.04, 22.47), n=1 | 0.78 (0.27, 2.22), n=2 | 1.15 (0.54, 2.46), n=3 |
|  | **Eta** | 0.45 (0.02, 8.79), n=1 | n=0 | 1.03 (0.04, 23.67), n=1 | Reference | 0.95 (0.05, 16.70), n=1 | 0.45 (0.02, 10.55), n=1 |
|  | **Omicron** | 1.20 (0.63, 2.28), n=3 | 0.88 (0.29, 2.64), n=3 | 1.29 (0.45, 3.69), n=2 | 1.05 (0.06, 18.45), n=1 | Reference | 1.05 (0.39, 2.79), n=3 |
|  | **Mixed** | 0.69 (0.33, 1.44), n=4 | 0.49 (0.16, 1.50), n=3 | 0.87 (0.41, 1.86), n=3 | 2.21 (0.09, 51.31), n=1 | 0.95 (0.36, 2.54), n=3 | Reference |
| **Preterm labor (COVID-19 onset <37w)** | **Pre-alpha** | Reference | 0.90 (0.57, 1.42), n=2 | 1.40 (0.75, 2.61), n=2 | n=0 | 0.99 (0.54, 1.82), n=2 | 0.90 (0.56, 1.44), n=2 |
|  | **Alpha** | 1.11 (0.70, 1.76), n=2 | Reference | 1.48 (0.84, 2.61), n=2 | n=0 | 1.06 (0.64, 1.75), n=2 | 1.00 (0.55, 1.81), n=2 |
|  | **Delta** | 0.71 (0.38, 1.33), n=2 | 0.68 (0.38, 1.19), n=2 | Reference | n=0 | 0.73 (0.46, 1.14), n=2 | 0.66 (0.37, 1.17), n=2 |
|  | **Omicron** | 1.01 (0.55, 1.85), n=2 | 0.94 (0.57, 1.55), n=2 | 1.38 (0.88, 2.17), n=2 | n=0 | Reference | 0.93 (0.56, 1.54), n=2 |
|  | **Mixed** | 1.11 (0.70, 1.78), n=2 | 1.00 (0.55, 1.81), n=2 | 1.51 (0.85, 2.68), n=2 | n=0 | 1.08 (0.65, 1.79), n=2 | Reference |
| **Hemorrhage** | **Pre-alpha** | Reference | 2.06 (1.08, 3.92), n=3 | 1.18 (0.57, 2.44), n=2 | 2.10 (0.87, 5.08), n=1 | 1.17 (0.71, 1.94), n=2 | 1.70 (1.04, 2.80), n=3 |
|  | **Alpha** | 0.49 (0.25, 0.93), n=3 | Reference | 0.55 (0.24, 1.26), n=2 | 1.04 (0.39, 2.78), n=1 | 0.59 (0.30, 1.13), n=2 | 0.82 (0.43, 1.56), n=3 |
|  | **Delta** | 0.85 (0.41, 1.76), n=2 | 1.82 (0.80, 4.14), n=2 | Reference | 1.76 (0.62, 4.99), n=1 | 1.08 (0.51, 2.30), n=1 | 1.35 (0.65, 2.81), n=2 |
|  | **Eta** | 0.48 (0.20, 1.15), n=1 | 0.96 (0.36, 2.57), n=1 | 0.57 (0.20, 1.61), n=1 | Reference | 0.61 (0.26, 1.47), n=1 | 0.78 (0.32, 1.87), n=1 |
|  | **Omicron** | 0.85 (0.52, 1.41), n=2 | 1.71 (0.89, 3.28), n=2 | 0.92 (0.43, 1.96), n=1 | 1.63 (0.68, 3.90), n=1 | Reference | 1.42 (0.87, 2.33), n=2 |
|  | **Mixed** | 0.59 (0.36, 0.97), n=3 | 1.22 (0.64, 2.33), n=3 | 0.74 (0.36, 1.55), n=2 | 1.29 (0.54, 3.10), n=1 | 0.70 (0.43, 1.15), n=2 | Reference |
| **Embolic disease** | **Pre-alpha** | Reference | 1.87 (0.08, 44.71), n=1 | 1.28 (0.05, 30.82), n=1 | 3.74 (0.16, 87.35), n=1 | 0.40 (0.02, 9.63), n=1 | 0.57 (0.02, 13.74), n=1 |
|  | **Alpha** | 0.53 (0.02, 12.76), n=1 | Reference | n=0 | n=0 | n=0 | n=0 |
|  | **Delta** | 0.78 (0.03, 18.79), n=1 | n=0 | Reference | n=0 | n=0 | n=0 |
|  | **Eta** | 0.27 (0.01, 6.23), n=1 | n=0 | n=0 | Reference | n=0 | n=0 |
|  | **Omicron** | 2.53 (0.10, 61.49), n=1 | n=0 | n=0 | n=0 | Reference | n=0 |
|  | **Mixed** | 1.77 (0.07, 42.90), n=1 | n=0 | n=0 | n=0 | n=0 | Reference |
| **Preeclampsia** | **Pre-alpha** | Reference | 1.31 (0.46, 3.70), n=3 | 1.13 (0.33, 3.90), n=2 | n=0 | 0.97 (0.44, 2.13), n=2 | 1.92 (0.77, 4.76), n=3 |
|  | **Alpha** | 0.77 (0.27, 2.17), n=3 | Reference | 0.65 (0.19, 2.18), n=2 | n=0 | 1.06 (0.33, 3.43), n=2 | 1.55 (0.50, 4.80), n=2 |
|  | **Delta** | 0.88 (0.26, 3.03), n=2 | 1.54 (0.46, 5.17), n=2 | Reference | n=0 | 1.49 (0.32, 6.90), n=1 | 2.04 (0.64, 6.52), n=2 |
|  | **Omicron** | 1.04 (0.47, 2.28), n=2 | 0.94 (0.29, 3.05), n=2 | 0.67 (0.15, 3.09), n=1 | n=0 | Reference | 1.61 (0.58, 4.43), n=2 |
|  | **Mixed** | 0.52 (0.21, 1.29), n=3 | 0.65 (0.21, 2.01), n=2 | 0.49 (0.15, 1.56), n=2 | n=0 | 0.62 (0.23, 1.71), n=2 | Reference |
| **Eclampsia** | **Pre-alpha** | Reference | n=0 | n=0 | n=0 | n=0 | n=0 |
|  | **Alpha** | n=0 | Reference | n=0 | n=0 | n=0 | n=0 |
|  | **Omicron** | n=0 | n=0 | n=0 | n=0 | Reference | n=0 |
|  | **Mixed** | n=0 | n=0 | n=0 | n=0 | n=0 | Reference |
| **Preeclampsia or eclampsia** | **Pre-alpha** | Reference | 1.48 (0.19, 11.42), n=2 | 0.77 (0.04, 15.67), n=1 | 2.25 (0.11, 44.35), n=1 | 0.91 (0.27, 3.10), n=2 | 0.72 (0.09, 5.72), n=2 |
|  | **Alpha** | 0.68 (0.09, 5.25), n=2 | Reference | n=0 | n=0 | 0.58 (0.08, 4.27), n=2 | n=0 |
|  | **Delta** | 1.30 (0.06, 26.54), n=1 | n=0 | Reference | n=0 | 1.54 (0.08, 31.48), n=1 | n=0 |
|  | **Eta** | 0.45 (0.02, 8.79), n=1 | n=0 | n=0 | Reference | 0.53 (0.03, 10.43), n=1 | n=0 |
|  | **Omicron** | 1.10 (0.32, 3.76), n=2 | 1.71 (0.23, 12.55), n=2 | 0.65 (0.03, 13.19), n=1 | 1.89 (0.10, 37.33), n=1 | Reference | 0.85 (0.11, 6.41), n=2 |
|  | **Mixed** | 1.39 (0.17, 11.07), n=2 | n=0 | n=0 | n=0 | 1.18 (0.16, 8.88), n=2 | Reference |
| **HDP (diagnosed at any time)** | **Pre-alpha** | Reference | 1.19 (0.40, 3.48), n=3 | 0.69 (0.27, 1.78), n=2 | 2.25 (0.11, 44.35), n=1 | 0.72 (0.27, 1.89), n=2 | 1.41 (0.57, 3.48), n=4 |
|  | **Alpha** | 0.84 (0.29, 2.48), n=3 | Reference | 0.56 (0.15, 2.06), n=1 | n=0 | 0.74 (0.10, 5.32), n=2 | 1.61 (0.46, 5.66), n=2 |
|  | **Delta** | 1.44 (0.56, 3.69), n=2 | 1.80 (0.49, 6.66), n=1 | Reference | n=0 | 1.54 (0.08, 31.48), n=1 | 2.56 (0.85, 7.71), n=1 |
|  | **Eta** | 0.45 (0.02, 8.79), n=1 | n=0 | n=0 | Reference | 0.53 (0.03, 10.43), n=1 | n=0 |
|  | **Omicron** | 1.39 (0.53, 3.68), n=2 | 1.35 (0.19, 9.69), n=2 | 0.65 (0.03, 13.19), n=1 | 1.89 (0.10, 37.33), n=1 | Reference | 0.68 (0.09, 5.01), n=2 |
|  | **Mixed** | 0.71 (0.29, 1.76), n=4 | 0.62 (0.18, 2.18), n=2 | 0.39 (0.13, 1.18), n=1 | n=0 | 1.48 (0.20, 10.94), n=2 | Reference |
| **HDP (diagnosed at or after COVID-19)** | **Pre-alpha** | Reference | 1.20 (0.38, 3.79), n=2 | 0.59 (0.21, 1.67), n=1 | n=0 | 0.53 (0.17, 1.68), n=1 | 1.00 (0.31, 3.19), n=2 |
|  | **Alpha** | 0.84 (0.26, 2.65), n=2 | Reference | 0.48 (0.13, 1.84), n=1 | n=0 | 0.50 (0.04, 6.88), n=1 | 0.85 (0.18, 4.00), n=1 |
|  | **Delta** | 1.69 (0.60, 4.75), n=1 | 2.09 (0.54, 7.99), n=1 | Reference | n=0 | n=0 | 1.78 (0.46, 6.86), n=1 |
|  | **Omicron** | 1.88 (0.60, 5.95), n=1 | 2.00 (0.15, 27.51), n=1 | n=0 | n=0 | Reference | 1.50 (0.10, 21.92), n=1 |
|  | **Mixed** | 1.00 (0.31, 3.21), n=2 | 1.17 (0.25, 5.49), n=1 | 0.56 (0.15, 2.16), n=1 | n=0 | 0.67 (0.05, 9.74), n=1 | Reference |
| **C-section** | **Pre-alpha** | Reference | 1.26 (1.03, 1.53), n=5 | 0.99 (0.80, 1.23), n=4 | 1.12 (0.48, 2.59), n=1 | 0.89 (0.76, 1.05), n=4 | 1.31 (1.10, 1.56), n=5 |
|  | **Alpha** | 0.80 (0.65, 0.97), n=5 | Reference | 0.85 (0.65, 1.12), n=4 | 0.76 (0.30, 1.89), n=1 | 0.66 (0.53, 0.81), n=4 | 1.05 (0.84, 1.31), n=5 |
|  | **Delta** | 1.01 (0.81, 1.24), n=4 | 1.17 (0.89, 1.54), n=4 | Reference | 0.75 (0.31, 1.81), n=1 | 0.86 (0.67, 1.10), n=3 | 1.11 (0.76, 1.63), n=4 |
|  | **Eta** | 0.89 (0.39, 2.07), n=1 | 1.32 (0.53, 3.29), n=1 | 1.33 (0.55, 3.21), n=1 | Reference | 0.89 (0.38, 2.06), n=1 | 0.95 (0.40, 2.25), n=1 |
|  | **Omicron** | 1.12 (0.95, 1.32), n=4 | 1.52 (1.23, 1.88), n=4 | 1.17 (0.91, 1.50), n=3 | 1.13 (0.48, 2.63), n=1 | Reference | 1.59 (1.23, 2.06), n=4 |
|  | **Mixed** | 0.76 (0.64, 0.91), n=5 | 0.95 (0.76, 1.19), n=5 | 0.90 (0.62, 1.32), n=4 | 1.05 (0.44, 2.48), n=1 | 0.63 (0.49, 0.81), n=4 | Reference |
| **Intrapartum C-section** | **Pre-alpha** | Reference | 1.46 (1.00, 2.14), n=5 | 1.08 (0.74, 1.57), n=4 | 0.65 (0.10, 4.48), n=1 | 0.94 (0.74, 1.18), n=4 | 1.23 (0.93, 1.63), n=5 |
|  | **Alpha** | 0.68 (0.47, 1.00), n=5 | Reference | 0.80 (0.53, 1.22), n=4 | 0.42 (0.05, 3.18), n=1 | 0.58 (0.38, 0.87), n=4 | 0.90 (0.64, 1.27), n=5 |
|  | **Delta** | 0.92 (0.64, 1.35), n=4 | 1.25 (0.82, 1.89), n=4 | Reference | 0.30 (0.04, 2.11), n=1 | 0.85 (0.56, 1.28), n=3 | 1.03 (0.70, 1.53), n=4 |
|  | **Eta** | 1.53 (0.22, 10.51), n=1 | 2.40 (0.31, 18.34), n=1 | 3.33 (0.47, 23.41), n=1 | Reference | 1.67 (0.24, 11.49), n=1 | 1.98 (0.28, 13.71), n=1 |
|  | **Omicron** | 1.07 (0.85, 1.35), n=4 | 1.74 (1.16, 2.61), n=4 | 1.18 (0.78, 1.78), n=3 | 0.60 (0.09, 4.11), n=1 | Reference | 1.38 (1.02, 1.87), n=4 |
|  | **Mixed** | 0.81 (0.61, 1.08), n=5 | 1.11 (0.79, 1.57), n=5 | 0.97 (0.65, 1.44), n=4 | 0.51 (0.07, 3.51), n=1 | 0.72 (0.53, 0.98), n=4 | Reference |
| **Maternal composite outcome** | **Pre-alpha** | Reference | 1.16 (0.86, 1.55), n=4 | 0.81 (0.50, 1.32), n=3 | 1.12 (0.55, 2.26), n=1 | 0.99 (0.77, 1.27), n=3 | 1.11 (0.81, 1.51), n=5 |
|  | **Alpha** | 0.86 (0.64, 1.16), n=4 | Reference | 0.90 (0.62, 1.32), n=3 | 0.87 (0.40, 1.90), n=1 | 0.55 (0.31, 0.98), n=3 | 0.97 (0.71, 1.32), n=5 |
|  | **Delta** | 1.23 (0.76, 2.00), n=3 | 1.11 (0.76, 1.61), n=3 | Reference | 0.81 (0.39, 1.70), n=1 | 0.75 (0.52, 1.09), n=2 | 1.05 (0.77, 1.42), n=3 |
|  | **Eta** | 0.89 (0.44, 1.80), n=1 | 1.15 (0.53, 2.52), n=1 | 1.23 (0.59, 2.58), n=1 | Reference | 1.00 (0.50, 2.02), n=1 | 1.07 (0.53, 2.18), n=1 |
|  | **Omicron** | 1.01 (0.79, 1.30), n=3 | 1.81 (1.02, 3.24), n=3 | 1.33 (0.92, 1.92), n=2 | 1.00 (0.49, 2.01), n=1 | Reference | 1.75 (0.97, 3.18), n=3 |
|  | **Mixed** | 0.90 (0.66, 1.24), n=5 | 1.04 (0.76, 1.41), n=5 | 0.96 (0.71, 1.29), n=3 | 0.93 (0.46, 1.90), n=1 | 0.57 (0.31, 1.03), n=3 | Reference |

HDP: Hypertensive disorders of pregnancy

C. **Fetal and Neonatal Morbidity & Mortality**

| **Outcome** | **Reference Strain** | **Pre-alpha** | **Alpha** | **Delta** | **Eta** | **Omicron** | **Mixed** |
| --- | --- | --- | --- | --- | --- | --- | --- |
| **Stillbirth** | **Pre-alpha** | Reference | 2.03 (0.27, 15.26), n=2 | 3.64 (0.99, 13.41), n=3 | n=0 | 1.39 (0.24, 8.02), n=3 | 3.54 (0.40, 31.20), n=2 |
|  | **Alpha** | 0.49 (0.07, 3.72), n=2 | Reference | 1.89 (0.38, 9.49), n=3 | n=0 | 0.71 (0.07, 7.43), n=2 | 2.36 (0.38, 14.84), n=2 |
|  | **Delta** | 0.27 (0.07, 1.01), n=3 | 0.53 (0.11, 2.66), n=3 | Reference | n=0 | 0.33 (0.06, 1.94), n=3 | 0.90 (0.25, 3.30), n=3 |
|  | **Eta** | n=0 | n=0 | n=0 | Reference | n=0 | n=0 |
|  | **Omicron** | 0.72 (0.12, 4.14), n=3 | 1.40 (0.13, 14.53), n=2 | 3.03 (0.51, 17.85), n=3 | n=0 | Reference | 2.51 (0.37, 17.17), n=3 |
|  | **Mixed** | 0.28 (0.03, 2.49), n=2 | 0.42 (0.07, 2.66), n=2 | 1.11 (0.30, 4.07), n=3 | n=0 | 0.40 (0.06, 2.72), n=3 | Reference |
| **Perinatal death** | **Pre-alpha** | Reference | 1.87 (0.28, 12.71), n=2 | 3.84 (0.89, 16.49), n=3 | n=0 | 2.81 (0.57, 13.85), n=2 | 5.16 (1.09, 24.37), n=2 |
|  | **Alpha** | 0.53 (0.08, 3.62), n=2 | Reference | 1.68 (0.26, 10.90), n=2 | n=0 | 1.30 (0.18, 9.57), n=2 | 2.88 (0.30, 27.21), n=1 |
|  | **Delta** | 0.26 (0.06, 1.12), n=3 | 0.60 (0.09, 3.86), n=2 | Reference | n=0 | 0.68 (0.08, 5.94), n=2 | 1.18 (0.26, 5.42), n=2 |
|  | **Omicron** | 0.36 (0.07, 1.76), n=2 | 0.77 (0.10, 5.65), n=2 | 1.48 (0.17, 12.92), n=2 | n=0 | Reference | 1.74 (0.15, 20.09), n=2 |
|  | **Mixed** | 0.19 (0.04, 0.92), n=2 | 0.35 (0.04, 3.28), n=1 | 0.85 (0.18, 3.90), n=2 | n=0 | 0.58 (0.05, 6.67), n=2 | Reference |
| **Early neonatal death** | **Pre-alpha** | Reference | 1.28 (0.13, 12.26), n=2 | 1.75 (0.28, 11.04), n=3 | n=0 | 2.32 (0.38, 14.16), n=2 | 1.41 (0.15, 13.49), n=2 |
|  | **Alpha** | 0.78 (0.08, 7.46), n=2 | Reference | 0.74 (0.03, 17.77), n=1 | n=0 | 1.66 (0.18, 14.94), n=2 | n=0 |
|  | **Delta** | 0.57 (0.09, 3.62), n=3 | 1.36 (0.06, 32.73), n=1 | Reference | n=0 | 1.41 (0.16, 12.71), n=2 | 1.16 (0.05, 28.11), n=1 |
|  | **Omicron** | 0.43 (0.07, 2.64), n=2 | 0.60 (0.07, 5.42), n=2 | 0.71 (0.08, 6.36), n=2 | n=0 | Reference | 0.67 (0.07, 6.00), n=2 |
|  | **Mixed** | 0.71 (0.07, 6.78), n=2 | n=0 | 0.86 (0.04, 20.77), n=1 | n=0 | 1.50 (0.17, 13.48), n=2 | Reference |
| **Neonatal death** | **Pre-alpha** | Reference | 1.28 (0.13, 12.26), n=2 | 2.81 (0.57, 13.84), n=4 | n=0 | 2.32 (0.38, 14.16), n=2 | 1.41 (0.15, 13.49), n=2 |
|  | **Alpha** | 0.78 (0.08, 7.46), n=2 | Reference | 1.21 (0.13, 11.40), n=2 | n=0 | 1.66 (0.18, 14.94), n=2 | n=0 |
|  | **Delta** | 0.36 (0.07, 1.75), n=4 | 0.83 (0.09, 7.78), n=2 | Reference | 0.97 (0.04, 22.47), n=1 | 0.61 (0.10, 3.70), n=3 | 0.41 (0.04, 3.92), n=2 |
|  | **Eta** | n=0 | n=0 | 1.03 (0.04, 23.67), n=1 | Reference | n=0 | n=0 |
|  | **Omicron** | 0.43 (0.07, 2.64), n=2 | 0.60 (0.07, 5.42), n=2 | 1.65 (0.27, 10.04), n=3 | n=0 | Reference | 0.67 (0.07, 6.00), n=2 |
|  | **Mixed** | 0.71 (0.07, 6.78), n=2 | n=0 | 2.42 (0.26, 22.96), n=2 | n=0 | 1.50 (0.17, 13.48), n=2 | Reference |
| **NICU** | **Pre-alpha** | Reference | 0.76 (0.10, 5.77), n=1 | 0.49 (0.06, 3.79), n=1 | 1.51 (0.21, 11.09), n=1 | 1.19 (0.46, 3.07), n=1 | 0.85 (0.26, 2.75), n=1 |
|  | **Alpha** | 1.32 (0.17, 10.12), n=1 | Reference | 0.65 (0.04, 9.88), n=1 | 2.00 (0.14, 29.28), n=1 | 1.57 (0.21, 12.01), n=1 | 1.13 (0.13, 9.64), n=1 |
|  | **Delta** | 2.04 (0.26, 15.81), n=1 | 1.54 (0.10, 23.49), n=1 | Reference | 3.08 (0.21, 45.62), n=1 | 2.43 (0.31, 18.77), n=1 | 1.74 (0.20, 15.05), n=1 |
|  | **Eta** | 0.66 (0.09, 4.86), n=1 | 0.50 (0.03, 7.32), n=1 | 0.32 (0.02, 4.80), n=1 | Reference | 0.79 (0.11, 5.77), n=1 | 0.56 (0.07, 4.64), n=1 |
|  | **Omicron** | 0.84 (0.33, 2.18), n=1 | 0.64 (0.08, 4.85), n=1 | 0.41 (0.05, 3.19), n=1 | 1.27 (0.17, 9.32), n=1 | Reference | 0.72 (0.22, 2.31), n=1 |
|  | **Mixed** | 1.17 (0.36, 3.78), n=1 | 0.89 (0.10, 7.56), n=1 | 0.57 (0.07, 4.97), n=1 | 1.77 (0.22, 14.55), n=1 | 1.39 (0.43, 4.48), n=1 | Reference |
| **Fetal composite outcome** | **Pre-alpha** | Reference | 0.95 (0.52, 1.74), n=5 | 0.80 (0.46, 1.38), n=3 | 0.50 (0.07, 3.41), n=1 | 0.41 (0.13, 1.28), n=3 | 1.16 (0.55, 2.45), n=5 |
|  | **Alpha** | 1.06 (0.58, 1.93), n=5 | Reference | 0.99 (0.61, 1.63), n=3 | 0.69 (0.08, 6.00), n=1 | 0.53 (0.15, 1.88), n=3 | 1.01 (0.60, 1.69), n=5 |
|  | **Delta** | 1.26 (0.72, 2.18), n=3 | 1.01 (0.61, 1.65), n=3 | Reference | 0.62 (0.08, 4.77), n=1 | 0.51 (0.14, 1.86), n=2 | 1.30 (0.75, 2.26), n=3 |
|  | **Eta** | 1.99 (0.29, 13.44), n=1 | 1.44 (0.17, 12.44), n=1 | 1.62 (0.21, 12.55), n=1 | Reference | 2.07 (0.30, 14.04), n=1 | 1.55 (0.22, 10.98), n=1 |
|  | **Omicron** | 2.42 (0.78, 7.54), n=3 | 1.89 (0.53, 6.73), n=3 | 1.98 (0.54, 7.27), n=2 | 0.48 (0.07, 3.29), n=1 | Reference | 1.76 (0.67, 4.67), n=3 |
|  | **Mixed** | 0.86 (0.41, 1.80), n=5 | 0.99 (0.59, 1.67), n=5 | 0.77 (0.44, 1.33), n=3 | 0.64 (0.09, 4.55), n=1 | 0.57 (0.21, 1.50), n=3 | Reference |

NICU: Neonatal intensive care unit

D. **Adverse Birth Outcomes**

| **Outcome** | **Reference Strain** | **Pre-alpha** | **Alpha** | **Beta** | **Delta** | **Epsilon** | **Eta** |
| --- | --- | --- | --- | --- | --- | --- | --- |
| **vLBW** | **Pre-alpha** | Reference | 1.26 (0.37, 4.37), n=3 | 4.15 (1.77, 9.72), n=4 | 3.74 (0.16, 87.35), n=1 | 1.40 (0.61, 3.22), n=4 | 1.87 (0.62, 5.61), n=4 |
|  | **Alpha** | 0.79 (0.23, 2.73), n=3 | Reference | 2.09 (0.54, 8.04), n=4 | n=0 | 0.84 (0.27, 2.57), n=4 | 1.69 (0.44, 6.51), n=3 |
|  | **Delta** | 0.24 (0.10, 0.57), n=4 | 0.48 (0.12, 1.84), n=4 | Reference | 0.97 (0.04, 22.47), n=1 | 0.44 (0.13, 1.50), n=3 | 0.63 (0.13, 3.01), n=4 |
|  | **Eta** | 0.27 (0.01, 6.23), n=1 | n=0 | 1.03 (0.04, 23.67), n=1 | Reference | 0.74 (0.04, 13.55), n=1 | n=0 |
|  | **Omicron** | 0.71 (0.31, 1.63), n=4 | 1.20 (0.39, 3.68), n=4 | 2.26 (0.66, 7.67), n=3 | 1.35 (0.07, 24.75), n=1 | Reference | 1.31 (0.45, 3.77), n=4 |
|  | **Mixed** | 0.53 (0.18, 1.60), n=4 | 0.59 (0.15, 2.27), n=3 | 1.60 (0.33, 7.70), n=4 | n=0 | 0.77 (0.27, 2.21), n=4 | Reference |
| **LBW** | **Pre-alpha** | Reference | 1.05 (0.69, 1.61), n=5 | 1.37 (0.90, 2.09), n=4 | 1.21 (0.17, 8.66), n=1 | 1.01 (0.75, 1.36), n=4 | 1.16 (0.78, 1.72), n=5 |
|  | **Alpha** | 0.95 (0.62, 1.45), n=5 | Reference | 1.25 (0.61, 2.56), n=4 | 2.00 (0.14, 29.28), n=1 | 0.97 (0.61, 1.55), n=4 | 1.16 (0.70, 1.91), n=5 |
|  | **Delta** | 0.73 (0.48, 1.11), n=4 | 0.80 (0.39, 1.65), n=4 | Reference | 1.03 (0.12, 8.98), n=1 | 0.90 (0.54, 1.51), n=3 | 0.97 (0.54, 1.71), n=4 |
|  | **Eta** | 0.83 (0.12, 5.93), n=1 | 0.50 (0.03, 7.32), n=1 | 0.97 (0.11, 8.50), n=1 | Reference | 1.67 (0.24, 11.49), n=1 | 0.56 (0.07, 4.64), n=1 |
|  | **Omicron** | 0.99 (0.73, 1.33), n=4 | 1.03 (0.65, 1.65), n=4 | 1.11 (0.66, 1.87), n=3 | 0.60 (0.09, 4.11), n=1 | Reference | 0.99 (0.51, 1.91), n=4 |
|  | **Mixed** | 0.86 (0.58, 1.28), n=5 | 0.87 (0.52, 1.43), n=5 | 1.04 (0.58, 1.84), n=4 | 1.77 (0.22, 14.55), n=1 | 1.01 (0.52, 1.97), n=4 | Reference |
| **eSGA** | **Pre-alpha** | Reference | 1.36 (0.59, 3.10), n=4 | 1.45 (0.63, 3.34), n=3 | n=0 | 0.37 (0.13, 1.02), n=3 | 1.38 (0.56, 3.40), n=4 |
|  | **Alpha** | 0.74 (0.32, 1.69), n=4 | Reference | 1.01 (0.32, 3.21), n=3 | n=0 | 0.34 (0.10, 1.18), n=1 | 0.93 (0.31, 2.76), n=3 |
|  | **Delta** | 0.69 (0.30, 1.60), n=3 | 0.99 (0.31, 3.14), n=3 | Reference | n=0 | 0.43 (0.10, 1.89), n=2 | 0.86 (0.29, 2.53), n=3 |
|  | **Omicron** | 2.70 (0.98, 7.41), n=3 | 2.94 (0.85, 10.21), n=1 | 2.30 (0.53, 9.99), n=2 | n=0 | Reference | 6.31 (0.66, 60.52), n=2 |
|  | **Mixed** | 0.73 (0.29, 1.79), n=4 | 1.08 (0.36, 3.19), n=3 | 1.16 (0.40, 3.40), n=3 | n=0 | 0.16 (0.02, 1.52), n=2 | Reference |
| **SGA** | **Pre-alpha** | Reference | 1.36 (0.65, 2.83), n=4 | 1.74 (0.90, 3.36), n=3 | n=0 | 0.58 (0.24, 1.38), n=3 | 2.09 (0.96, 4.53), n=4 |
|  | **Alpha** | 0.73 (0.35, 1.53), n=4 | Reference | 1.18 (0.48, 2.88), n=3 | n=0 | 0.37 (0.12, 1.13), n=2 | 1.41 (0.58, 3.41), n=3 |
|  | **Delta** | 0.58 (0.30, 1.11), n=3 | 0.85 (0.35, 2.08), n=3 | Reference | n=0 | 0.43 (0.10, 1.89), n=2 | 1.41 (0.74, 2.71), n=3 |
|  | **Omicron** | 1.73 (0.72, 4.13), n=3 | 2.74 (0.89, 8.43), n=2 | 2.30 (0.53, 9.99), n=2 | n=0 | Reference | 2.59 (0.84, 8.00), n=2 |
|  | **Mixed** | 0.48 (0.22, 1.04), n=4 | 0.71 (0.29, 1.71), n=3 | 0.71 (0.37, 1.36), n=3 | n=0 | 0.39 (0.12, 1.20), n=2 | Reference |
| **vPTB** | **Pre-alpha** | Reference | 1.53 (0.77, 3.05), n=5 | 3.49 (2.05, 5.93), n=4 | 6.04 (0.59, 61.92), n=1 | 1.07 (0.60, 1.89), n=4 | 1.68 (0.85, 3.32), n=5 |
|  | **Alpha** | 0.65 (0.33, 1.30), n=5 | Reference | 2.08 (0.99, 4.38), n=4 | 2.08 (0.14, 30.54), n=1 | 0.66 (0.32, 1.38), n=4 | 1.11 (0.49, 2.51), n=4 |
|  | **Delta** | 0.29 (0.17, 0.49), n=4 | 0.48 (0.23, 1.01), n=4 | Reference | 3.08 (0.21, 45.62), n=1 | 0.35 (0.16, 0.77), n=3 | 0.60 (0.24, 1.48), n=4 |
|  | **Eta** | 0.17 (0.02, 1.70), n=1 | 0.48 (0.03, 7.04), n=1 | 0.32 (0.02, 4.80), n=1 | Reference | 0.49 (0.06, 3.87), n=1 | 0.28 (0.03, 2.88), n=1 |
|  | **Omicron** | 0.93 (0.53, 1.65), n=4 | 1.51 (0.73, 3.14), n=4 | 2.83 (1.30, 6.15), n=3 | 2.03 (0.26, 16.01), n=1 | Reference | 1.40 (0.69, 2.86), n=4 |
|  | **Mixed** | 0.60 (0.30, 1.18), n=5 | 0.90 (0.40, 2.05), n=4 | 1.68 (0.68, 4.15), n=4 | 3.54 (0.35, 36.15), n=1 | 0.71 (0.35, 1.45), n=4 | Reference |
| **vPTB (COVID-19 onset <34w)** | **Pre-alpha** | Reference | n=0 | n=0 | n=0 | n=0 | n=0 |
|  | **Alpha** | n=0 | Reference | n=0 | n=0 | n=0 | n=0 |
|  | **Delta** | n=0 | n=0 | Reference | n=0 | n=0 | n=0 |
|  | **Omicron** | n=0 | n=0 | n=0 | n=0 | Reference | n=0 |
|  | **Mixed** | n=0 | n=0 | n=0 | n=0 | n=0 | Reference |
| **PTB** | **Pre-alpha** | Reference | 0.88 (0.56, 1.37), n=5 | 1.48 (0.85, 2.60), n=4 | 1.51 (0.21, 11.09), n=1 | 1.06 (0.77, 1.44), n=4 | 1.09 (0.71, 1.68), n=5 |
|  | **Alpha** | 1.14 (0.73, 1.79), n=5 | Reference | 1.59 (0.92, 2.74), n=4 | 2.08 (0.14, 30.54), n=1 | 1.12 (0.69, 1.81), n=4 | 1.09 (0.60, 1.97), n=4 |
|  | **Delta** | 0.67 (0.38, 1.18), n=4 | 0.63 (0.37, 1.09), n=4 | Reference | 1.54 (0.15, 15.54), n=1 | 0.74 (0.48, 1.14), n=3 | 0.84 (0.47, 1.50), n=4 |
|  | **Eta** | 0.66 (0.09, 4.86), n=1 | 0.48 (0.03, 7.04), n=1 | 0.65 (0.06, 6.54), n=1 | Reference | 1.08 (0.15, 7.68), n=1 | 1.13 (0.15, 8.25), n=1 |
|  | **Omicron** | 0.95 (0.69, 1.29), n=4 | 0.89 (0.55, 1.45), n=4 | 1.35 (0.88, 2.08), n=3 | 0.92 (0.13, 6.56), n=1 | Reference | 0.87 (0.55, 1.38), n=4 |
|  | **Mixed** | 0.92 (0.60, 1.41), n=5 | 0.92 (0.51, 1.67), n=4 | 1.19 (0.67, 2.14), n=4 | 0.89 (0.12, 6.47), n=1 | 1.15 (0.72, 1.82), n=4 | Reference |
| **PTB (COVID-19 onset <37w)** | **Pre-alpha** | Reference | 0.89 (0.55, 1.45), n=2 | 1.57 (0.81, 3.06), n=2 | n=0 | 1.08 (0.65, 1.78), n=2 | 0.75 (0.44, 1.30), n=2 |
|  | **Alpha** | 1.12 (0.69, 1.83), n=2 | Reference | 1.63 (0.90, 2.96), n=2 | n=0 | 1.12 (0.66, 1.89), n=2 | 0.84 (0.43, 1.64), n=2 |
|  | **Delta** | 0.64 (0.33, 1.24), n=2 | 0.61 (0.34, 1.11), n=2 | Reference | n=0 | 0.68 (0.43, 1.08), n=2 | 0.51 (0.27, 0.97), n=2 |
|  | **Omicron** | 0.93 (0.56, 1.54), n=2 | 0.89 (0.53, 1.51), n=2 | 1.46 (0.92, 2.31), n=2 | n=0 | Reference | 0.74 (0.42, 1.32), n=2 |
|  | **Mixed** | 1.33 (0.77, 2.29), n=2 | 1.19 (0.61, 2.31), n=2 | 1.96 (1.03, 3.73), n=2 | n=0 | 1.35 (0.76, 2.40), n=2 | Reference |

vLBW: Very low birthweight; LBW: Low birthweight; eSGA: Extremely small-for-gestational age; SGA: Small-for-gestational age; vPTB: Very Preterm birth; PTB: Preterm birth
